# Cervicovaginal immune mediators increase when young women begin to have sexual intercourse: a prospective study and meta-analysis

**DOI:** 10.1101/2022.03.31.22273275

**Authors:** Sean M. Hughes, Claire N. Levy, Fernanda L. Calienes, Katie A. Martinez, Stacy Selke, Kenneth Tapia, Bhavna H. Chohan, Lynda Oluoch, Catherine Kiptinness, Anna Wald, Mimi Ghosh, Liselotte Hardy, Kenneth Ngure, Nelly R. Mugo, Florian Hladik, Alison C. Roxby

## Abstract

**Background:** Adolescent girls and young women (AGYW) are at high risk of sexually transmitted infections (STIs). It is unknown whether beginning to have sexual intercourse causes changes to immune mediators in the cervicovaginal tract that contribute to this risk.

**Methods:** We collected cervicovaginal lavages from Kenyan AGYW in the months before and after first penile-vaginal sexual intercourse and measured the concentrations of 20 immune mediators. We compared concentrations pre- and post-first sex using mixed effects models. Secondary analyses included adjustment for possible confounding factors. We additionally performed a systematic review to identify similar studies and combined them with our results by meta-analysis of individual participant data.

**Results:** We included 180 samples from 95 AGYW, with 44% providing only pre-first sex samples, 35% matched pre and post, and 21% only post. We consistently detected 19/20 immune mediators, all of which increased post-first sex (median increase 54%; p<0.05 for 13/19; Holm-Bonferroni-adjusted p<0.05 for IL-1β, IL-2 and CXCL8). Effects remained similar after adjusting for confounding factors including STIs and Nugent score.

Our systematic review identified two eligible studies, one of 93 Belgian participants and the other of 18 American participants. Nine immune mediators were measured in at least 2/3 studies. Meta-analysis confirmed higher levels post-first sex for 8/9 immune mediators (median increase 47%; p<0.05 for six mediators, most prominently IL-1α, IL-1β and CXCL8).

**Conclusions:** Cervicovaginal immune mediator concentrations increased after the beginning of sexual activity independently of confounding factors including STIs. Results were consistent across three studies conducted on three different continents.

## Introduction

Adolescent girls and young women (AGYW) aged 15-24 are at high risk for sexually transmitted infections (STIs) and are disproportionately affected by human immunodeficiency virus (HIV), accounting for as many as 80% of new HIV infections in some countries.^1-3^ In Kenya in particular, a quarter of new HIV infections occur in AGYW.^4^ Because of the disproportionate impact of HIV on AGYW, understanding the behavioral and physiological components of heightened STI risk in this population presents an opportunity to reduce HIV infections.

The period following first penile-vaginal sexual intercourse (“first sex”) marks the start of vulnerability to STIs and is associated with higher acquisition of bacterial vaginosis and STIs than later in life. The reasons for this increased susceptibility remain unclear, because (1) studies of mucosal immunity in AGYW are challenging, and (2) it can be difficult to distinguish immune changes that are a consequence of STI acquisition from changes that occur independently of STIs. Understanding changes in cervicovaginal tract (CVT) immune mediators following first sex may help identify interventions to decrease the risk of STI and HIV acquisition in AGYW.

In this study, we measured immune mediators in cervicovaginal lavage (CVL) specimens collected from a unique longitudinal cohort of Kenyan AGYW,^1,2^ an especially vulnerable population. By comparing specimens collected in the months before and after first sex, our goal was to measure changes in CVT immune mediators following start of sexual intercourse. Extensive clinical information was available about participants, allowing adjustment for potential confounding factors, including acquisition of STIs.

To generalize our findings to broader populations, we additionally conducted a systematic review of published literature to identify other studies of cervicovaginal immune mediators in AGYW before and after first sexual intercourse. We sought individual participant data from study authors and performed meta-analyses of individual immune mediators, assessing changes in immune mediator concentration before and after first sex.

## Results

### Characteristics of cohort

We selected 195 samples for this study. We excluded 15 samples due to unavailable covariate information (such as total protein or PSA), extensive blood contamination, and presence of rare infections (*Trichomonas vaginalis*, *Neisseria gonorrhoeae*, genital HSV-1 or HSV-2 DNA; **Figure 1A**). The final sample set included 180 samples from 95 participants.

**Figure 1.**
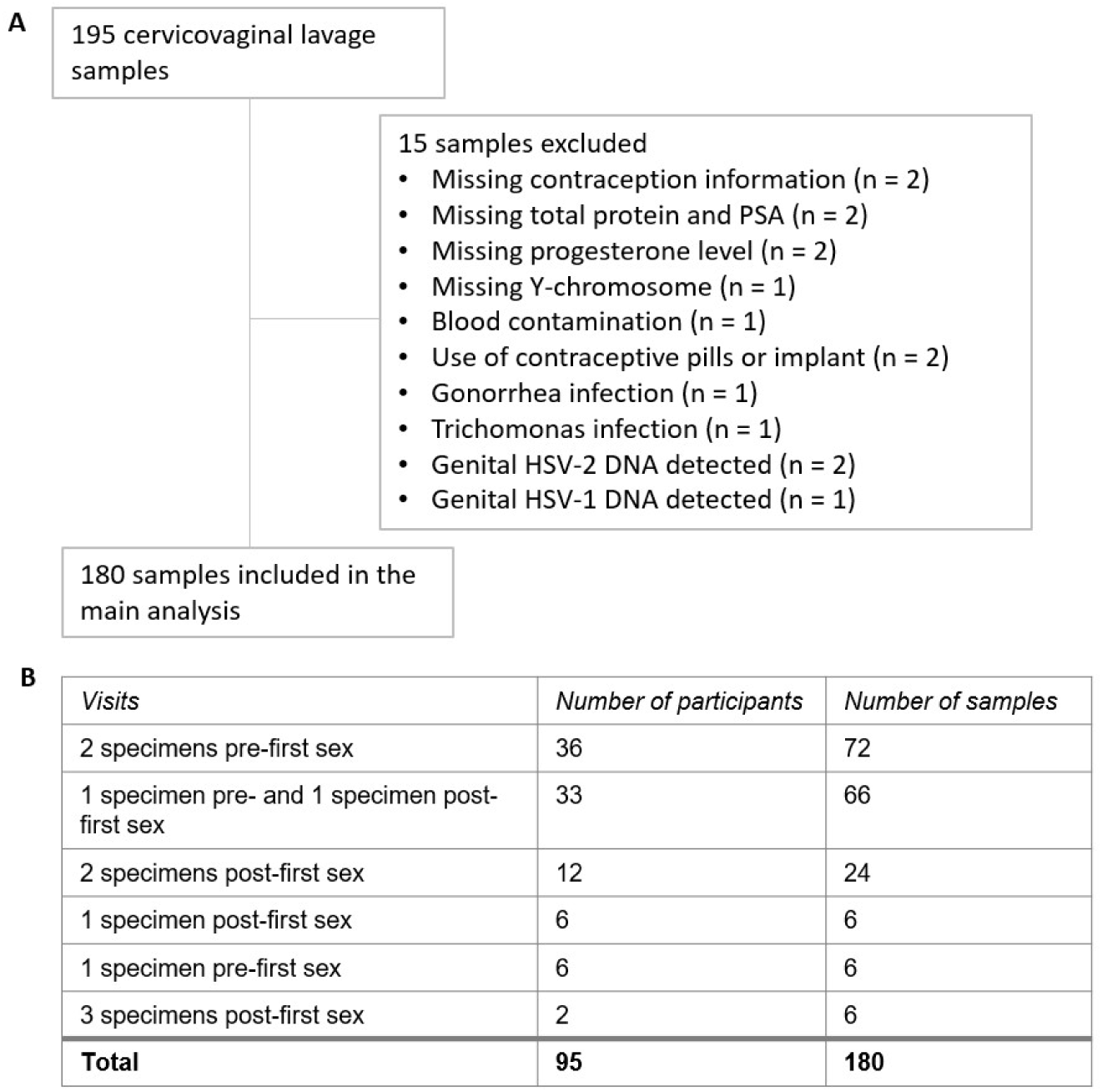
Cervicovaginal lavage sample selection. **A** Cervicovaginal lavage sample selection, collected from adolescent girls and young women in a longitudinal cohort study. **B** Timepoints for collection of cervicovaginal lavage specimens pre and post-first sexual intercourse.

Of these, 111 samples from 75 participants were classified as pre-first sex and 57 samples from 45 participants were obtained after reported first sex. In addition, 12 samples from 8 participants were classified as post-sex due to presence of Y-chromosome DNA, PSA or because of pregnancy or a positive *Chlamydia trachomatis* (CT) test at that visit or a prior visit. Pre-first sex samples were collected a median of 344 days before first sex (IQR 72-687), while post-first sex samples were collected a median of 66 days (27-105) after. We captured matched pre- and post-first sex specimens from 33 participants (66 specimens); the remaining participants provided either only pre-first sex specimens (42 participants; 78 specimens) or only post-first sex specimens (20 participants; 36 specimens; **Figure 1B**).

The median age of participants in the final sample set was 19.1 years (IQR 18.1-19.6). Post-first sex samples came from participants who were, on average, older and more likely to have BV (**Table 1**). By definition, only the post-first sex group included samples from pregnant participants, participants using contraception, and samples where Y-chromosome, PSA, CT, or HSV-2 were detected.

**Table 1.** Comparison of social, demographic and biological characteristics of adolescent girls and young women at timepoints of cervicovaginal lavage specimens selected for the study. Each participant can appear in this table more than once.

|  | Pre-first sex<br>timepoints<br>N = 111 specimens<sup>a</sup> | Post-first sex<br>timepoints<br>N = 69 specimens<sup>a</sup> | p-value |
| --- | --- | --- | --- |
| Age (years) | 18.80 (17.80, 19.45) | 19.30 (18.60, 19.80) | <0.001<sup>b</sup> |
| Menstrual phase |  |  |  |
| Luteal | 39 (35%) | 26 (38%) | 0.73<sup>c</sup> |
| Follicular | 64 (58%) | 35 (51%) | 0.36<sup>c</sup> |
| Other | 8 (7.2%) | 8 (12%) | 0.21<sup>c</sup> |
| Pregnant |  |  | –<sup>d</sup> |
| Negative | 111 (100%) | 64 (93%) |  |
| Positive | 0 (0%) | 5 (7.2%) |  |
| Contraception |  |  | –<sup>d</sup> |
| None | 111 (100%) | 54 (78%) |  |
| Emergency pills | 0 (0%) | 4 (5.8%) |  |
| Condoms | 0 (0%) | 11 (16%) |  |
| Y-chromosome |  |  | –<sup>d</sup> |
| Negative | 111 (100%) | 55 (80%) |  |
| Positive | 0 (0%) | 14 (20%) |  |
| Prostate specific antigen |  |  | –<sup>d</sup> |
| Negative | 111 (100%) | 67 (97%) |  |
| Positive | 0 (0%) | 2 (2.9%) |  |
| Bacterial vaginosis |  |  |  |
| Negative (Nugent score 0-3) | 102 (92%) | 54 (78%) | 0.08<sup>c</sup> |
| Intermediate (4-6) | 6 (5.4%) | 6 (8.7%) | 0.82<sup>c</sup> |
| Positive (7-10) | 3 (2.7%) | 9 (13%) | 0.02<sup>c</sup> |
| Chlamydia |  |  | –<sup>d</sup> |
| Negative | 111 (100%) | 59 (86%) |  |
| Positive | 0 (0%) | 10 (14%) |  |
| HSV-1 |  |  | >0.9<sup>c</sup> |
| Seronegative | 4 (3.6%) | 2 (2.9%) |  |
| Seropositive | 107 (96%) | 67 (97%) |  |
| HSV-2 |  |  | –<sup>d</sup> |
| Seronegative | 111 (100%) | 66 (96%) |  |
| Seropositive | 0 (0%) | 3 (4.3%) |  |

<sup>a</sup> Statistics presented: median (IQR) or n (%)
<sup>b</sup> Linear mixed effects regression
<sup>c</sup> Logistic mixed effects regression with binary combinations, comparing each category to all other samples (for example, luteal compared to pooled follicular and other).
<sup>d</sup> Not tested for difference between groups because this variable can only be positive at post-first sex time points by definition.

### Immune mediator measurements pre- and post-first sex

Concentrations of 20 immune mediators were measured **(Figure 2**). IL-1RA was detected at the highest concentrations. Sixteen immune mediators were detectable in most specimens. Four were detected in fewer than half of the samples: IFN-γ (27%), IL-10 (23%), IL-12p70 (20%), and IFN-α2A (7%); due to lack of detection, IFN-α2A was excluded from further analysis.

**Figure 2.**
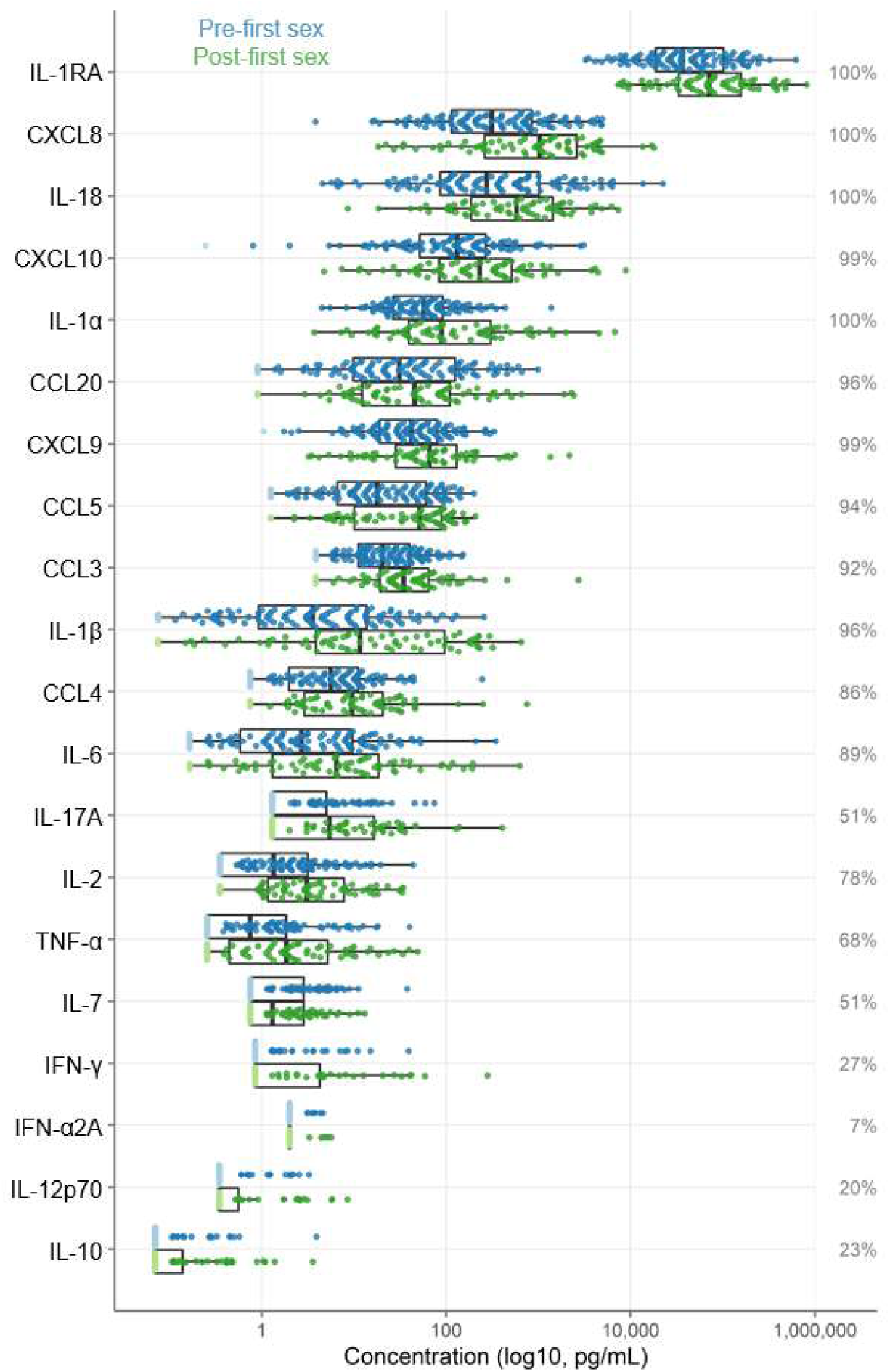
Immune factor concentrations from cervicovaginal lavage specimens from adolescent girls and young women, comparing specimens from before and after first sexual intercourse. Blue indicates specimens prior to first sex and green indicates samples after first sex. Specimens below the limit of detection are indicated as lighter colors and were set to half the limit of detection. Percentages indicate the percent of samples within the detectable range of the assays.

As shown in **Figure 2**, immune mediator concentrations were higher after first sex. We quantified the differences between pre- and post-first sex samples (**Figure 2**). In all cases, the concentration (**Figure 3A, Table 2**) or proportion detectable (**Figure 3B, Table 3**) was higher. The differences were significant at p<0.05 for most (13/19) immune mediators and remained significant for IL-1β, IL-2 and CXCL8 after adjustment for multiple comparisons (adjustment for 19 immune mediators).

**Figure 3.**
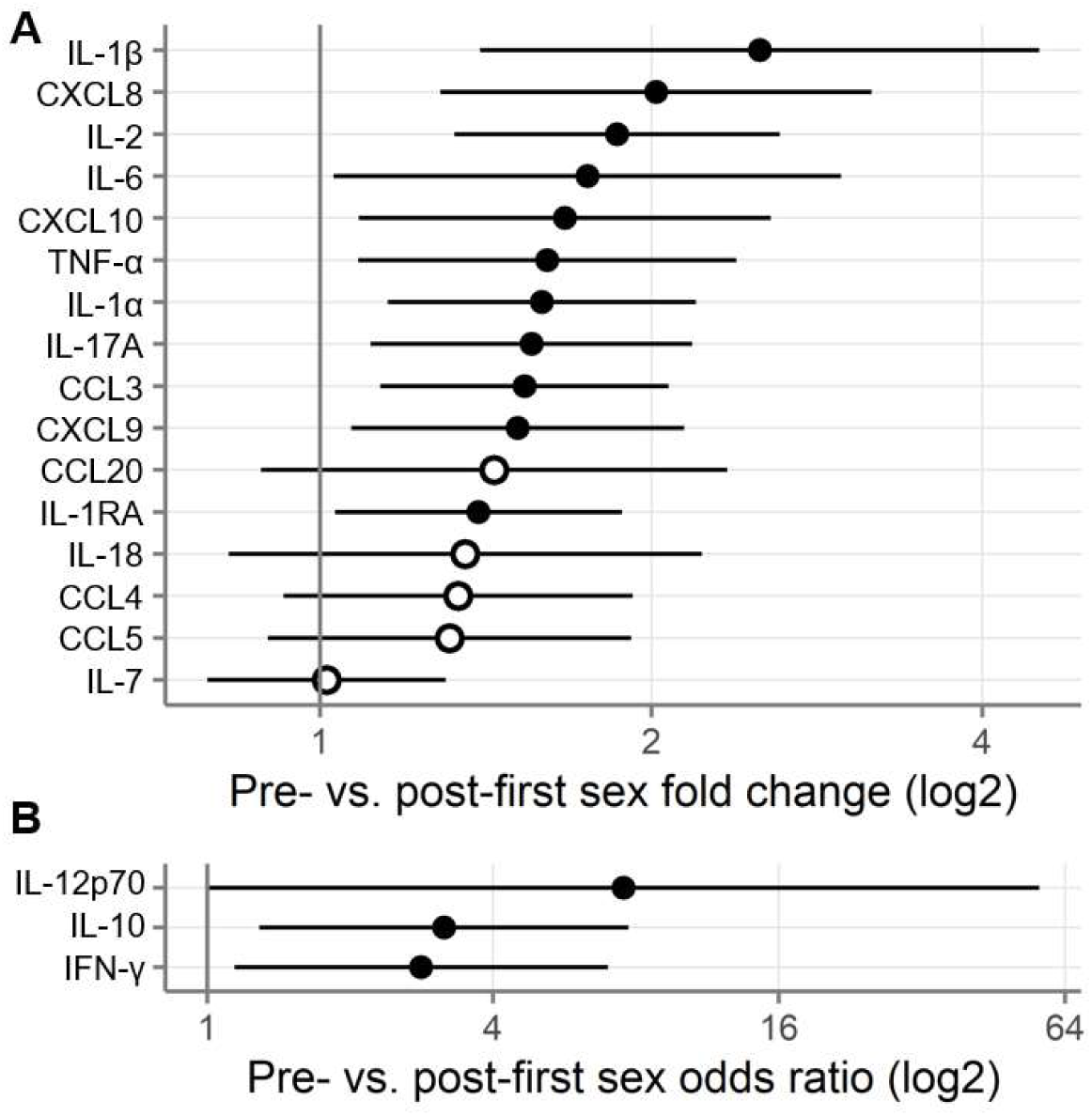
Comparison of immune mediators in cervicovaginal lavage samples pre- and post-first sexual intercourse. Univariate mixed-effect models with first sex as fixed effect and participant as random effects. Symbols indicate the mean and horizontal lines indicate the 95% confidence intervals. Filled symbols indicate p<0.05 while open symbols indicate p≥0.05. Vertical lines at 1 indicate no difference between pre- and post-first sex. **A** Log2-fold change between pre- and post-first sex; positive numbers indicate greater quantities of immune mediators post-first sex. **B** Odds ratio (log2) comparing the odds of the immune mediator being detected above the lower limit of detection (for immune mediators detected in fewer than half of the samples). Positive numbers indicate greater detection of immune mediators post-first sex.

**Table 2.** Comparison of immune mediators in cervicovaginal lavage samples pre- and post-first sexual intercourse. Results of univariate mixed-effect models with first sex as fixed effect and participant as random effects. Mean log2 difference values above 0 indicate higher concentrations post-first sex. Adjusted p-values are adjusted by Holm-Bonferroni for 19 immune mediators.

| Immune mediator | Mean log2 difference | Standard error | P-value | Adjusted p-value |
| --- | --- | --- | --- | --- |
| IL-2 | 0.90 | 0.25 | 4.7E-4 | 0.009 |
| IL-1β | 1.33 | 0.43 | 0.002 | 0.044 |
| CXCL8 | 1.01 | 0.33 | 0.003 | 0.045 |
| IL-1α | 0.67 | 0.24 | 0.006 | 0.088 |
| CCL3 | 0.62 | 0.22 | 0.006 | 0.091 |
| IL-17A | 0.64 | 0.25 | 0.011 | 0.154 |
| TNF-α | 0.69 | 0.29 | 0.020 | 0.236 |
| CXCL10 | 0.74 | 0.32 | 0.021 | 0.236 |
| CXCL9 | 0.60 | 0.26 | 0.021 | 0.236 |
| IL-1RA | 0.48 | 0.22 | 0.032 | 0.258 |
| IL-6 | 0.81 | 0.39 | 0.041 | 0.284 |
| CCL4 | 0.42 | 0.27 | 0.124 | 0.621 |
| CCL20 | 0.53 | 0.36 | 0.146 | 0.621 |
| CCL5 | 0.39 | 0.28 | 0.166 | 0.621 |
| IL-18 | 0.44 | 0.36 | 0.232 | 0.621 |
| IL-7 | 0.02 | 0.18 | 0.921 | 0.921 |

**Table 3.** Comparison of immune mediators in cervicovaginal lavage samples pre- and post-first sexual intercourse. Results of univariate mixed-effect models with first sex as fixed effect and participant as random effects. Mean log2 difference values above 0 indicate that the immune mediator was more often detected post-first sex. Adjusted p-values are adjusted by Holm-Bonferroni for 19 immune mediators.

| Immune mediator | Mean log2 difference | Standard error | P-value | Adjusted p-value |
| --- | --- | --- | --- | --- |
| IL-10 | 1.15 | 0.46 | 0.012 | 0.154 |
| IFN-γ | 1.04 | 0.46 | 0.025 | 0.236 |
| IL-12p70 | 2.02 | 1.03 | 0.050 | 0.300 |

### Immune mediator concentrations over time

We next wondered whether immune mediator concentrations increased all at once at first sex or cumulatively over time. We therefore assessed immune mediator concentrations relative to date of first sex. Dates were available for 80 pre-first sex samples from 59 participants and 60 post-first sex samples from 49 participants.

As shown in **Figure 4**, immune mediator concentrations were generally stable for three years prior to first sex and then increased sharply in the year following first sex. This pattern is consistent with cumulative increases in immune mediator concentrations following first sex. By contrast, for a stepwise increase, the expected pattern would be stable high concentrations following first sex with a flat slope; this pattern was not observed.

**Figure 4.**
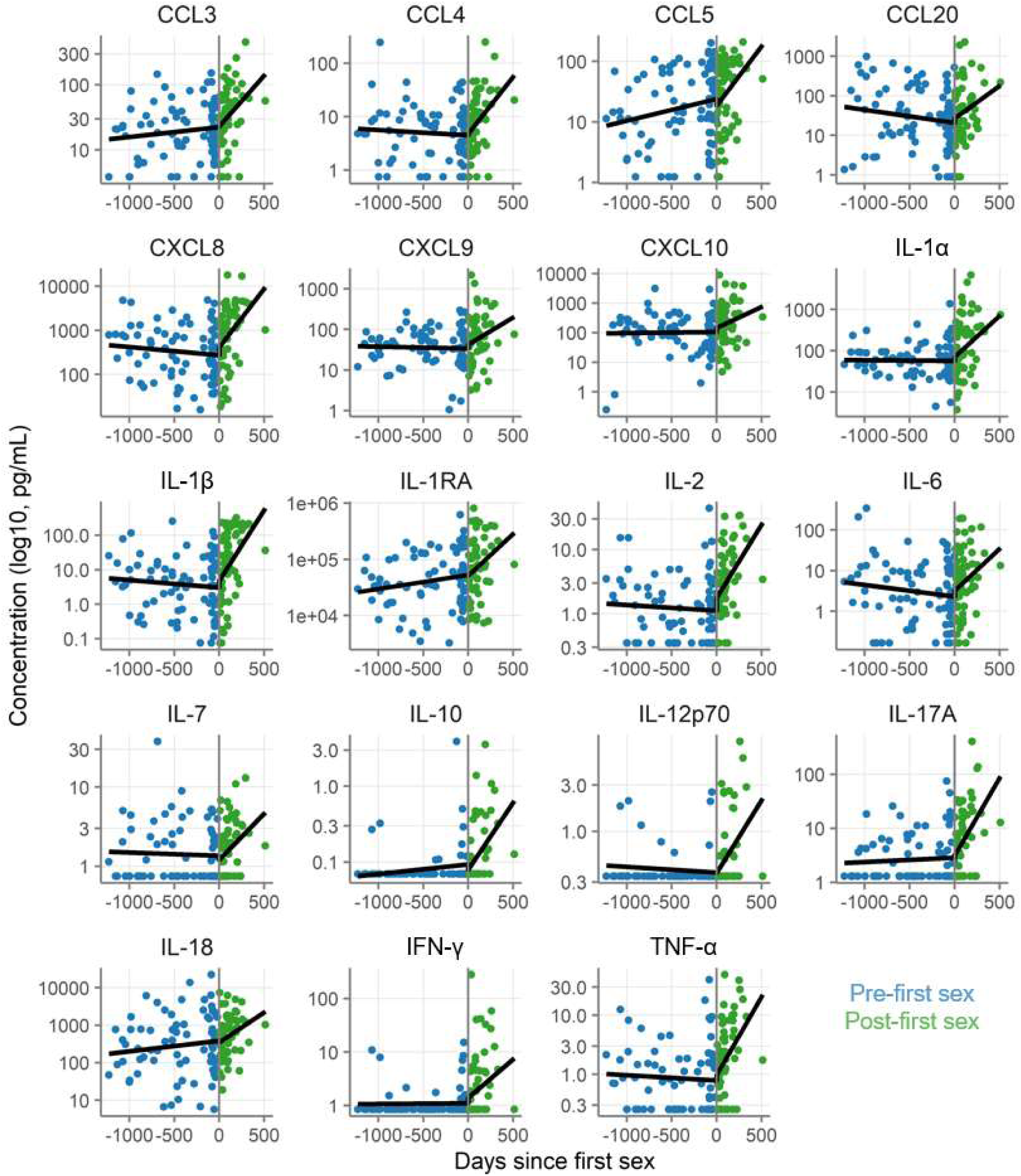
Concentration of immune mediators in cervicovaginal lavage samples relative to day of first sex. Each symbol depicts the concentration in a single sample. Black lines show the slopes from univariate mixed-effect models with days since first sex as fixed effect and participant as random effects, fit separately for pre- and post-first sex samples.

### Assessment of confounding

We next sought to determine whether the differences between pre- and post-first sex could be explained by co-variates that differed between the groups or that are known to affect immune mediator concentrations (**Table 1**). Specifically, we assessed whether the effect sizes and directions remained similar after adjusting for age, menstrual phase, pregnancy, contraception, Nugent score, CT infection, and HSV-2 seropositivity. As shown in **Figure 5AB** and **Supplemental File 2**, the effect sizes from these multivariate models were similar to those from univariate models, indicating that the co-variates did not explain the differences observed between pre- and post-first sex samples. The correlation between the effect sizes from the primary and multivariate analysis was strong (**Fig 5C**, top; Pearson r = 0.71, p = 0.002).

**Figure 5.**
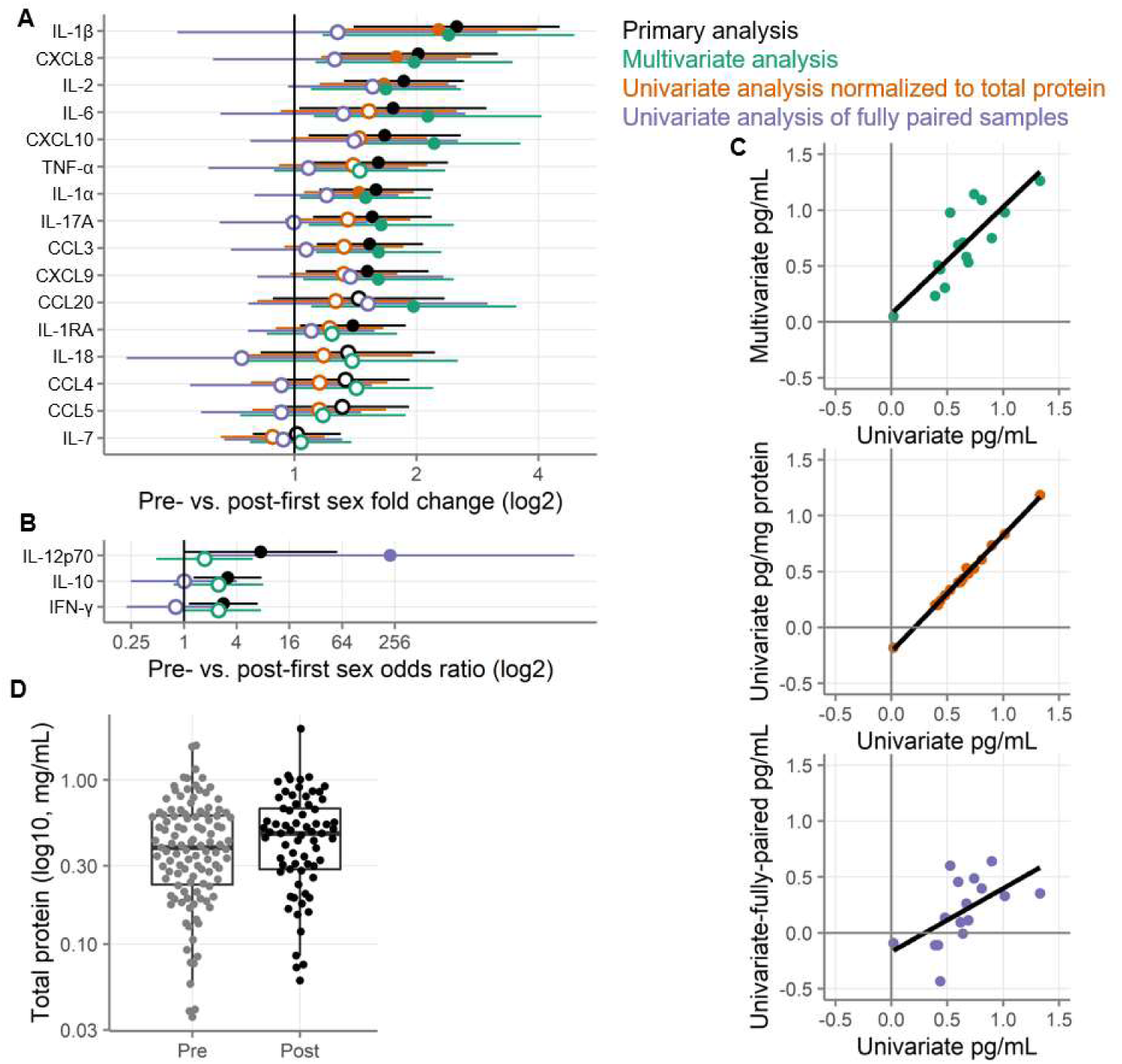
Alternative analysis strategies for the association of first sexual intercourse and quantities of cervicovaginal immune mediators. Filled symbols indicate p-value < 0.05 and open symbols indicate p-values ≥ 0.05. Vertical lines at 1 indicate no difference between pre- and post-first sex. Black symbols show the same primary analysis as Figure 3. Green symbols show a multivariate analysis adjusted for age, menstrual phase, pregnancy, contraception, Nugent score, Chlamydia infection, and HSV-2 seropositivity. Orange symbols show the primary univariate analysis performed on concentrations normalized to total protein concentrations (pg/mg protein). Purple symbols show the primary analysis repeated on only those samples from participants who provided both pre- and post-first sex samples. A Log2-fold change between pre- and post-first sex, where positive numbers indicate higher values post-sex. B Odds ratio (log2) for the immune mediator being detected above the lower limit of detection, where positive numbers indicate higher value post-sex. Symbols indicate the mean and horizontal lines indicate the 95% confidence intervals. C Total protein concentrations in cervicovaginal lavage specimens. D Correlation of effect sizes from the primary analysis (x-axis) with the three alternative analyses (y-axis). Each point represents the log2 difference between pre- and post-first sex (as shown in A) in concentrations for a single immune mediator.

Two of the largest differences between univariate and multivariate analysis were for CXCL10 and CCL20, where the multivariate analysis estimated much larger effect sizes. This difference is explained by Nugent score: high Nugent scores were more common post-first sex and were associated with strong negative effects on CXCL10 and CCL20.

To determine whether the pre- and post-first sex differences were driven by differences in protein recovery, we adjusted the immune mediator concentrations by total protein concentrations. This analysis modestly reduced the effect size estimates between pre- and post-first sex (**Figure 5AB**). This result is consistent with the slightly higher total protein concentrations observed in post-first sex CVLs (**Figure 5D**, 0.25 log2 fold change mg/mL, p=0.12). In all cases except for IL-7, the immune mediator concentrations remained higher post-first sex after normalization to total protein. Though the effect sizes were lower after normalization to total protein, they were extremely highly correlated with the primary analysis (**Fig 5C**, middle; Pearson r = 0.998, p = 2.7E-18).

### Fully paired analysis

As described above, only a minority of participants provided both pre- and post-first sex samples; about two thirds of participants provided only pre-first sex or only post-first sex samples. This is a consequence of the difficulty of capturing participants at the precise right moment in their lives and is consistent with prior studies.^5,6^

We repeated our univariate analysis on the 66 samples from the 33 participants who provided paired pre- and post-first sex samples. As shown in **Figure 5AB**, immune mediator concentrations were generally higher post-first sex in this subset of samples, but the effects were smaller and failed to reach statistical significance. The effect sizes from the fully paired analysis had a medium correlation with the effect sizes from the full data set (**Fig 5C**, Pearson r = 0.56, p = 0.02).

The specimens from the fully-paired subset were obtained sooner after reported date of first sex than the specimens from the participants who only provided post specimens (p = 4.7E-8). In the fully-paired subset, the post-first sex samples came only about a month after first sex (median 31.5 days). In contrast, the post-first sex samples from the other participants came about four months after first sex (median 157 days). The age of the participants at the time of first sex, however, was similar (18.9 years in each subset, p = 0.90).

### Systematic review and meta-analysis of immune mediator changes post-first sex

We performed a systematic review to identify comparable studies. Of 147 abstracts retrieved through our search, 7 were assessed as potentially eligible and 2 studies were determined to be eligible after review of the full-texts. We obtained individual participant data from both studies. The first was a cross-sectional study by ELISA of 11-19 year olds in Washington DC with 18 CVL samples from 10 participants post-first sex and 8 participants pre-first sex. This study found a statistically significant decrease in TNF-α, while most other mediators were higher (but not statistically significant) post-first sex.^7^ The other study longitudinally followed Belgian 14-19 year olds, with 269 swab samples from 93 participants, 9 of whom provided both pre- and post-first sex samples, 43 of whom provided only pre samples, and 41 of whom provided only post samples. Using Luminex, this study found increases in IL-1α, IL-1β, and CXCL8 post-first sex.^6^ No important issues were identified in checking the integrity of the individual participant data received for these two studies. Risk of bias was low (scores of 7/7 for ^6^ and the study reported here) to moderate (score of 4/7 for ^7^). IPD was obtained from all eligible studies, so there is no additional risk of bias from missing data.

Including our study reported here, 9 immune mediators were measured in at least 2 studies and were eligible for meta-analysis (**Figure 6**; **Table 4**). Meta-analyses identified six immune mediators as having higher concentrations post-first sex (IL-1α, IL-1β, IL-6, CXCL8, CCL4, and CCL5, all p<0.05; all p<0.05 after adjustment for multiple comparisons except for IL-6 and CCL4 [adjusted p = 0.066]). The remaining three remaining immune mediators all had meta-analysis p>0.05, with two higher post-first sex (CXCL10 and CCL20) and TNF-α showing the opposite. Supplemental File 6 contains detailed plots for each immune mediator included in the meta-analyses: the concentration data from every sample for each study and a forest plot showing each study’s effect size and weighting for each immune mediator.

**Figure 6.**
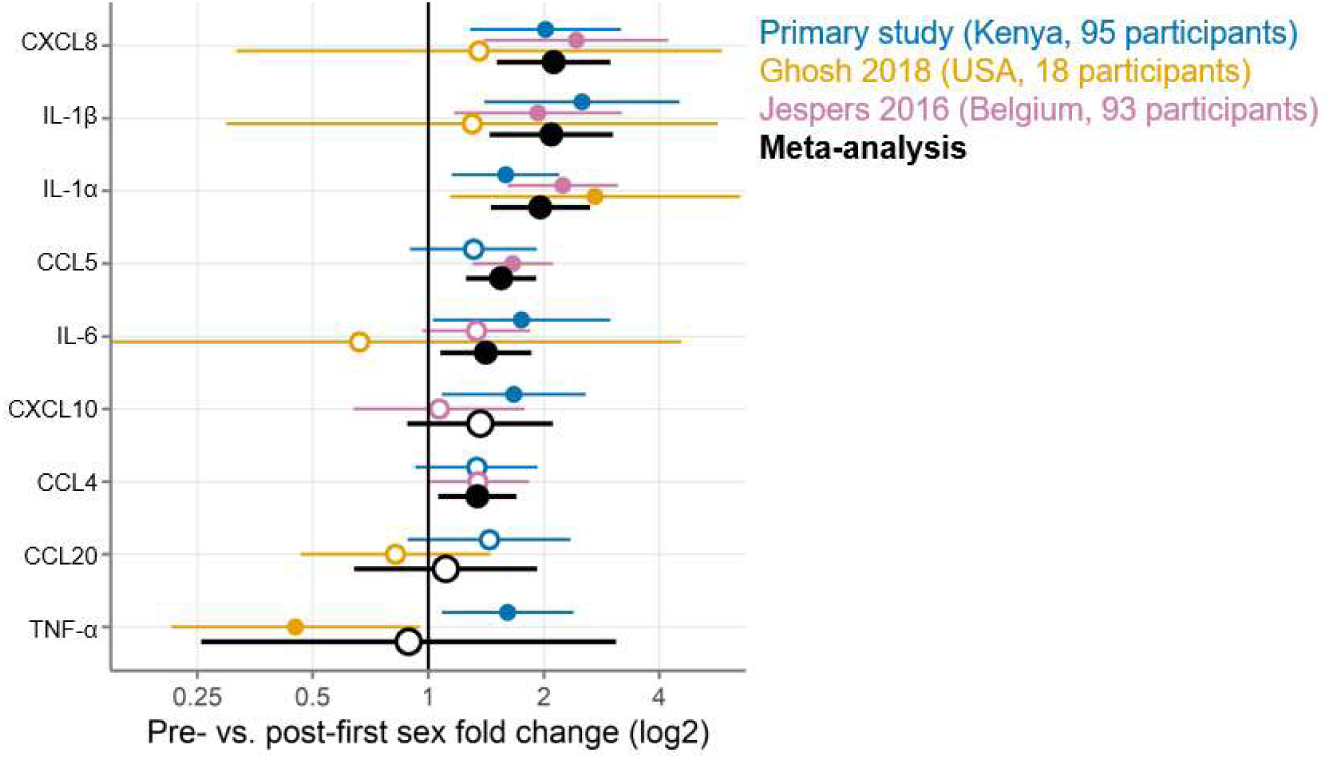
Meta-analysis comparing 3 studies measuring cervicovaginal immune mediators in adolescent girls and young women before and after first sexual intercourse. Log2-fold change in immune mediator concentrations between pre- and post-first sexual intercourse; positive numbers indicate higher values post-first sex. Symbols indicate the mean and horizontal lines indicate the 95% confidence intervals. Filled symbols indicate p-value < 0.05 while open symbols indicate p-values ≥ 0.05. Vertical lines at 1 indicate no difference between pre- and post-first sex. Colors indicate the source of the data (blue, primary study; gold, Ghosh 2018; pink, Jespers 2016; black, meta-analysis).

**Table 4.** Meta-analyses of immune mediator concentrations in cervicovaginal samples comparing pre- and post-first sexual intercourse. Results of random effects meta-analyses using inverse variance pooling. Mean log2 difference values above 0 indicate higher concentrations post-first sex. Adjusted p-values are adjusted by Holm-Bonferroni for 9 immune mediators.

| Immune mediator | Mean log2 difference | Standard error | P-value | Adjusted p-value | Number of studies | I<sup>2</sup> |
| --- | --- | --- | --- | --- | --- | --- |
| IL-1α | 0.97 | 0.22 | 1.0E-5 | 9.1E-5 | 3 | 0.28 |
| CXCL8 | 1.08 | 0.25 | 1.5E-5 | 1.2E-4 | 3 | 0 |
| CCL5 | 0.63 | 0.16 | 5.2E-5 | 3.6E-4 | 2 | 0.05 |
| IL-1β | 1.06 | 0.27 | 9.3E-5 | 5.6E-4 | 3 | 0 |
| IL-6 | 0.50 | 0.20 | 0.013 | 0.066 | 3 | 0 |
| CCL4 | 0.42 | 0.17 | 0.014 | 0.066 | 2 | 0 |
| CXCL10 | 0.45 | 0.32 | 0.164 | 0.492 | 2 | 0.42 |
| CCL20 | 0.15 | 0.40 | 0.712 | 1 | 2 | 0.54 |
| TNF-α | -0.17 | 0.92 | 0.851 | 1 | 2 | 0.89 |

We used statistical heterogeneity (I^2^) to assess the comparability of the three studies included in the meta-analysis, with an I^2^ of 0% indicating that all between-study variation can be explained by random sampling and higher values indicating clinical, biological, or methodological diversity between studies.^8^ In our meta-analysis, statistical heterogeneity was generally low (I^2^ of 0% for IL-1β, IL-6, CXCL8, and CCL4) to moderate (I^2^ of 5-55% for IL-1α, CCL5, CXCL10, and CCL20; **Table 4**). TNF-α had high statistical heterogeneity between studies (I^2^ = 88.5%).

## Discussion

In this study of a large cohort of Kenyan AGYW with specimens collected pre- and post-first sex, we found increased levels of CVT immune mediators post-first sex. The strongest evidence was for IL-1β, IL-2, and CXCL8, but all measured immune mediators followed a similar pattern. These differences were not explained by cofactors including BV and STIs, which are known causes of CVT inflammation. These results were robustly tested with several analytical approaches: limiting to only paired samples, adjusting for cofactors, and adjusting for multiple comparisons, but with all of these methods, the increased levels of immune mediators post-sex remained consistent.

We further performed a systematic review and meta-analysis of individual participant data, identifying two previous studies conducted in Belgium and the US.^6,7^ Meta-analysis combining all three studies confirmed increased concentrations post-first sex for 8/9 immune mediators, with particularly strong evidence for IL-1α, IL-1β, IL-6, and CXCL8. The impressive agreement across these three studies strengthens the conclusion that a wide range of CVT immune mediators increase following first sex.

While it has long been suspected that sexual activity induces CVT changes, our study, with post-first sex samples collected a median of only 66 days after first sex, clarifies that inflammatory changes occur very early in sexual activity. In fact, the rate of increase for the median immune mediator was about 0.006 log2 pg/mL per day (Fig 4), suggesting a doubling in concentration within about four months. Further, these changes appear to occur even without any incipient BV or STI, and might need to be considered a normal component of sexual activity. These inflammatory changes may be important clinically in modifying risk of STI acquisition and in fertility.

There has been speculation that AGYW are at increased risk of STI due to physiological differences in the CVT compared to older women, in addition to behavioral risk factors.^9-12^ Our research shows that after AGYW start to have sex, most immune mediators increase in concentration. Whether the increase in immune mediators brings protection from or vulnerability to STIs is unclear. Increased inflammation may prepare the CVT to prevent infection upon exposure. At the same time, increased expression of inflammatory mediators may recruit CD4 T cells, increasing the abundance of HIV target cells and potentially the risk of HIV infection.

In addition to preventing infection, immune mediators play important roles in implantation and pregnancy. Therefore, the increases in immune mediators we observed post-first sex may be relevant for fertility. For example, IL-6 induces sperm capacitation, increasing its fertilizing ability.^13^ In addition, trophoblast cells secrete immune mediators to attract and regulate immune cells within the placenta, which is a necessary process for successful pregnancy.^14^

### Mechanism of immune mediator increase

Several mechanisms may play a role in the association of sexual activity with increased CVT immune mediator concentrations: BV, semen exposure, vaginal practices including washing, and physical microtrauma.

Jespers et al. reported that the increased CVT cytokines they observed post-sex were largely mediated by increases in *G. vaginalis* and *A. vaginae*.^6^ Numerous studies show that BV is rare prior to first sex and increases thereafter.^15-22^ In addition, BV is associated with recent sexual activity, especially exposure to semen.^23-27^ BV is associated with increased CVT IL-1α and IL-1β, which agrees with our finding of higher IL-1α and IL-1β post-first sex. However, BV is also associated with reduced CXCL10,^28^ which conflicts with our finding of increased CXCL10 after first sex. Further, the increases in immune mediators we observed post-first sex remained after adjustment for Nugent score. Thus, changes to vaginal microbiota alone cannot explain the immune mediator changes we observed.

Vaginal washing changes the microbiome and increases BV.^29,30^ In this cohort, vaginal washing was reported by about 30% of participants, but data was not available for vaginal washing for the particular specimens analyzed here. However, because we adjusted for Nugent score, we were able to adjust for vaginal washing practices disruptive enough to change microbiome composition.

Exposure to semen has been associated with increases in cervical leukocytes^31-33^ and inflammatory cytokines, especially IL-6 and CXCL10, in most^25,26,33-36^ but not all studies.^37,38^ *In vitro* exposure to seminal plasma induces cytokines including IL-1α, IL-6, and CXCL8 in cervical and vaginal epithelial cells^39^ and cervical explants.^40-42^ Others have suggested that increases in immune mediators following exposure to semen may be a result of semen directly transferring cytokines into the CVT.^43^ However, detection of immune mediators directly transferred by semen is unlikely to explain our results, because semen was rarely detected in our study (by Y-chromosome DNA and PSA).

Penile-vaginal sexual intercourse can cause microtrauma in the vaginal walls,^44^ and this process may be inflammatory. However, exposure to semen may be necessary for the inflammatory response to intercourse, because condoms have been shown to block that response.^33^ Our study was unable to evaluate the role of microtrauma in the inflammatory response observed.

### Strengths and limitations

The strengths of our study include a large, well-characterized cohort with multiple specimens per participant, as well as the robustness of our findings to adjustment for possible confounding factors. In addition, our systematic review and meta-analysis places our results in the context of what is known in the literature and synthesizes all three studies. Despite the diversity of these studies and populations, all three studies showed consistent results.

An important limitation of our study is possible misclassification of specimens from pre-sexual activity. It is important to note that all three cohorts presented here were specifically designed to assess the sexual activity of adolescents and used best practices in ascertaining this challenging key variable. Of the 123 samples where participants reported no sexual activity, we classified 12 (9.8%) as post-first sex based on STI, pregnancy, Y-chromosome, or PSA results. In addition, false positive test results could have resulted in misclassification of pre-sex samples as post-sex. Since misclassification would be most likely for pre-sex specimens, misclassification would reduce any differences between pre- and post-sex specimens. In other words, if we were able to correct for any misclassification, this would likely increase the observed differences. Therefore, any residual misclassification remaining in our dataset would be unlikely to change our conclusions.

In a secondary analysis, we adjusted for a number of possibly confounding factors, including Nugent score and STIs. The results of these models were little changed from the results of our primary, univariate analysis. It could be argued that Nugent score and STIs are along the causal pathway and might cause the effects we observed, rather than confound them, in which case adjusting for them would spuriously reduce the effects we observed. However, the biological relationships are not well-understood, and Nugent score and STIs are both known to affect CVT immune mediators in other contexts, so we chose to adjust for them. Ultimately, the conclusion we draw from this analysis is that the results we observed are robust to a variety of alternative analytical approaches.

Given the consistency of results across three different studies, our results are likely generalizable to most AGYW. Our study did include a number of cases of chlamydia and BV, increasing its generalizability, given the commonness of these conditions. However, our study notably included few cases of HSV-2, a very common STI, which may reduce its generalizability if HSV-2 modifies the effect of sexual activity on immune mediators. Similarly, we excluded a small number of samples due to trichomonas or gonorrhea infection or active local HSV-1/HSV-2 production because there were too few samples to reliably analyze.

We observed higher total protein concentrations in the post-first sex samples, translating to smaller differences between pre- and post-first sex groups when immune mediator concentrations were normalized to total protein. However, immune mediators remained higher post-first sex even after normalizing to total protein, so global protein upregulation does not completely explain this phenomenon.

Smaller increases in immune markers were seen in the fully paired subset. A possible explanation for the smaller effect is that immune mediator concentrations increase over time post-first sex (Figure 4). The post-first sex samples in the fully paired subset were collected sooner after first sex (median 32 days) than those from the participants who provided only post-first sex samples (median 157 days). Thus, the effects may have been weaker in the fully paired subset because the samples were obtained such a short time after first sex.

The three studies included in the meta-analysis differed in important ways which could affect comparability. These differences included country of origin, presence of BV in the population, sample collection method,^45^ and immunoassay platform.^46^ However, statistical heterogeneity was low to moderate for most immune mediators, indicating that the between-study variability was largely what would be expected due to random sampling. The high statistical heterogeneity for TNF-α indicates a need for further study of that mediator. Because we obtained the raw individual participant data from each study, we were able to analyze all three studies using the same methods, which improves their comparability.

### Conclusions

We identified consistent increases in immune mediators when comparing vaginal specimens collected pre- and post-first penile-vaginal sexual intercourse. The dynamic nature of the inflammatory milieu after sexual activity may be a catalyst for changes that could promote acquisition of STIs. Because these specimens are from AGYW with little preexisting BV or STI, our research indicates that this inflammatory milieu may provide risk independent of STIs or vaginal dysbiosis. Further research should focus on the exact causes of inflammation associated with first sex and whether this potentially harmful inflammation also offers any benefits. We envision that this information will contribute to expanding our toolset in the fight against the high STI rates observed in AGYW.

## Methods

### Clinical cohort and study procedures

This study used specimens collected from the Kenya Girls Study, a previously described longitudinal cohort study of AGYW^1,2^. Briefly, AGYW aged 16-20 were recruited in Thika, Kenya. Enrolled participants returned quarterly, where they were interviewed about their sexual behavior and provided vaginal swabs, CVLs, and blood. For the sub-study described in this paper, we selected a group of 195 samples from the larger Kenya girls study, including samples from participants who provided only pre-first sex samples, participants who provided only post-first sex samples, and participants who provided both pre- and post-first sex samples. The sample size was chosen based on selection of all available samples from participants who reported sexual intercourse and a matching number of samples from participants who did not.

Human subjects approval was obtained from the Kenya Medical Research Institute Scientific Ethics Review Unit (protocol 2760) and the University of Washington Institutional Review Board (number 00000946). Participants under age 18 provided written informed assent and written informed consent was obtained from a parent/guardian. Participants assented privately from parents/guardians, after asking questions and deciding whether they wanted to participate free of parental influence. Participants age 18 or older provided written informed consent.

CVLs were collected using flexible tubing in the vagina to avoid a speculum examination. 5 cc of sterile saline was instilled into the vagina via tubing by the study clinician, left for 15 seconds, then aspirated into the syringe. Lavage fluid was spun for 10 minutes at 800×g; supernatant was removed, re-spun for 10 minutes, and stored in 2 mL aliquots at -80°C.

Participants were tested for STIs and bacterial vaginosis (BV). Vaginal swabs were tested for *Neisseria gonorrhea* (NG), *Chlamydia trachomatis* (CT), and *Trichomonas vaginalis* (TV) using the Gen-Probe APTIMA test (Hologic, Marlborough, MA) and for HSV-1 and HSV-2 by in-house PCR. Nugent scoring was performed on smears from vaginal swabs for BV; scores ≥7 were considered BV, scores 4-6 were considered intermediate, and scores ≤3 were considered no dysbiosis.^47^. Blood was tested for HIV using the Vironostika HIV Uni-Form II Ag-Ab (Biomerieux, Marcy-l’Etoile, France) and for HSV-1 and HSV-2 antibodies using in-house Western blot.

STI tests were performed annually in the parent study. Therefore, STI testing had not been performed at some timepoints selected for this sub-study. For samples with no concurrent STI result, we determined STI status as follows: If the annual tests both before and after the untested visit were negative, we inferred a negative result for that visit. If either annual test was positive, STI testing was performed for that timepoint. Negative CT and NG tests were inferred for 28 samples as well as negative TV tests for 45 samples.

Participants were tested for pregnancy via rapid urinary pregnancy test if they reported missed menses. Serum samples were tested for progesterone by automated immunoassay (Cobas E411, Roche, Basel, Switzerland). Participants were defined to be in the follicular phase if serum progesterone was <3 ng/mL, in the luteal phase if serum progesterone was ≥3 ng/mL, and ‘other’ if more than 35 days since the start of their last menstrual period had passed (indicating irregular menstruation, use of hormonal contraception, or pregnancy). Participants were assumed not to be using contraception until they reported sexual intercourse.

Prostate-specific antigen (PSA) was measured in CVL using the Human Kallikrein 3/PSA DuoSet ELISA (R&D Systems, Minneapolis, MN). Samples were considered positive if the PSA concentration was ≥10 ng/mL. Total protein concentrations were measured the Pierce BCA Protein Assay Kit (Thermo Fisher Scientific, Waltham, MA). PSA and total protein concentrations were calculated using four-parameter logistic curves fit to the standard curves. Vaginal swabs were tested for Y-chromosome DNA by Quantifiler® Duo DNA Quantification Kit (Life Technologies).

### Immune mediator quantification

We selected 20 immune mediators to measure in CVLs. These immune mediators were selected based on prior human and/or non-human primate studies as being consistently up- or down-regulated prior to HIV or STI transmission events, and therefore playing a key role in genital tract immunity to sexually transmitted infections. Concentrations of immune mediators were measured in CVL using Meso Scale Discovery (MSD) R-Plex/U-Plex kits according to the manufacturer’s instructions, and read on the MESO QuickPlex SQ 120. Pre- and post-first sex samples were present on every plate and the scientists were blinded to sample identity. CVLs were diluted 100-fold for measurement of IL-1RA and 10-fold for CXCL9 and CCL5. Concentrations were determined using four parameter logistic fits in MSD Discovery Workbench software.

### Definition of first sexual intercourse

Samples were categorized as pre- or post-first sex based on participant report of ever engaging in penile-vaginal penetrative sex. In addition, visits were categorized as post-first sex if at that visit or a previous visit, the participant was pregnant, Y-chromosome DNA was detected in a vaginal swab sample, PSA was detected in a CVL, or a vaginal swab tested positive for NG, CT, or TV.

### Statistical analysis

Data analysis was conducted using R^48^ version 4.0.0 with the packages plater^49^ and tidyverse^50^ in addition to the statistical packages described below.

For MSD, concentrations of each analyte were averaged across replicate wells and log2- transformed. Replicates with concentrations below the reported lower limits of detection were assigned the value of half the lower limit of detection. Samples were defined as detectable if the concentration was above the lower limit of detection for at least one of the two replicate wells. Some samples were excluded due to rare STIs or missing data (fully described in Figure 1), these exclusion criteria were not pre-established.

We performed primary and secondary analyses. For the primary analysis, we used univariate models to estimate the difference between cytokine concentrations in pre- and post-first sex samples (assessed by effect size and p-value). Pre- and post-first sex were modeled as a fixed effect and participant as a random effect to account for multiple samples per participant using mixed effect models with the R packages lme4^51^ and lmerTest.^52^ Log2-transformed immune mediator concentrations were used as the outcome for immune mediators if more than 50% of samples were above the limit of detection. For the remaining immune mediators, logistic models were used with the outcome being above/below the lower limit of detection. We adjusted for multiple comparisons by the Holm-Bonferroni method.^53^

As secondary analyses, we used multivariate models to assess whether those differences were explained by other factors known to affect immune mediator concentrations in CVLs (assessed by a substantial reduction in effect size compared to the univariate model). We fit multivariate models with additional fixed effects: age, menstrual phase, pregnancy, contraception, Nugent score, CT, and HSV-2 antibody status. In addition, we repeated our univariate models using immune mediator concentrations normalized to total protein.

All code and raw data are available in Supplemental File 1. All results are available in Supplemental File 2.

### Systematic review and meta-analysis

We designed our systematic review and meta-analysis in compliance with the Preferred Reporting Items for a Systematic Review and Meta-Analysis of Individual Participant Data (PRISMA-IPD) guidelines.^54^ The PRISMA-IPD checklist is included as Supplemental File 3.

#### Study eligibility criteria

Studies were eligible if they reported original immunoassay data on immune mediator concentrations in CVT fluid samples from AGYW before and after first vaginal sexual intercourse. Immunoassay data included any antibody-based methods (such as ELISA, bead-based assays, and electrochemiluminescence assays). CVT fluid samples could be collected by CVL, menstrual cup, or swab. Each study was required to include samples from before and after first penile-vaginal sexual intercourse, but we did not require that there be pre- and post-first sex samples from each participant. Samples were required to be categorized as pre- or post-first sex based on participant report of ever engaging in penile-vaginal penetrative sex. In addition, visits were categorized as post-first sex based on pregnancy, detection of sperm antigen or DNA, or a positive STI test.

#### Study identification and inclusion

We searched PubMed on August 28, 2021 for articles published in English using the search strategy in Supplemental File 4. Abstracts from search results were reviewed independently by two reviewers (SMH and CNL) using abstrackr.^55^ If either reviewer judged the study to be potentially eligible based on the abstract, then full-text articles were obtained and reviewed by both reviewers. Study inclusion was determined by consensus between the two reviewers.

#### Data collection and verification

We sought individual participant data via email with the authors of eligible studies. For each sample, we collected the participant identifier, the pre/post-first sex status of the sample, and the immune mediator concentrations in pg/mL. For each study, we collected the type of immunoassay used, the type of sample collected, and the country of sample origin. Individual participant data were verified by replicating analyses published in the prior manuscripts. Pre/post-first sex status was standardized across studies: in particular, one study included samples from participants reporting genital touching without penile-vaginal sexual intercourse. We categorized these samples as pre-first sex in accordance with our definition above.

#### Data analysis

The outcome of interest was the difference in immune mediator concentrations between pre- and post-first sex samples. We performed meta-analyses on all immune mediators present in at least two included studies. We used a two-stage approach for meta-analysis. First, we reanalyzed the individual participant data from the eligible studies. Studies with multiple samples per participant were analyzed using mixed effects models as above; studies with single samples per participant were analyzed using simple linear models. For all studies, immune mediator concentrations (pg/mL) were log2-transformed. In the second stage, we performed random-effects meta-analysis using the inverse-variance method in the R package meta.^56^ We quantified heterogeneity using I^2^. We did not attempt to explore variation in effects by study-level characteristics due to the low number of studies available.

#### Risk of bias

We assessed risk of bias within each study using a modified Newcastle-Ottawa Quality Assessment scale (Supplemental File 5), ranging from 0 (high risk of bias) to 7 (low risk of bias).

## Data Availability

All raw data are available in Supplemental File 1

## Acknowledgments

The authors would like to thank the following for their contributions: study staff and participants in Thika, Kenya; the Endocrine Technologies Core (NIH P51OD011092) at the Oregon National Primate Research Center for measuring progesterone concentrations; Dr Scott McClelland and the University of Washington/University of Nairobi East Africa STI Laboratory in Mombasa, Kenya for STI testing; Diana Louden from the University of Washington Health Sciences Library for devising the systematic review search strategy; Dr. Veronica Gomez-Lobo for performing the clinical components of the Washington DC study.

## Supplemental files

Supplemental File 1. R code and raw data to reproduce the analysis.

Supplemental File 2. Results of analyses.

Supplemental File 3. PRISMA-IPD Checklist.

Supplemental File 4. Systematic review search terms.

Supplemental File 5. Risk of bias assessment scale.

Supplemental File 6. Plots of every meta-analysis separately.

### Supplemental File 3 - PRISMA-IPD Checklist

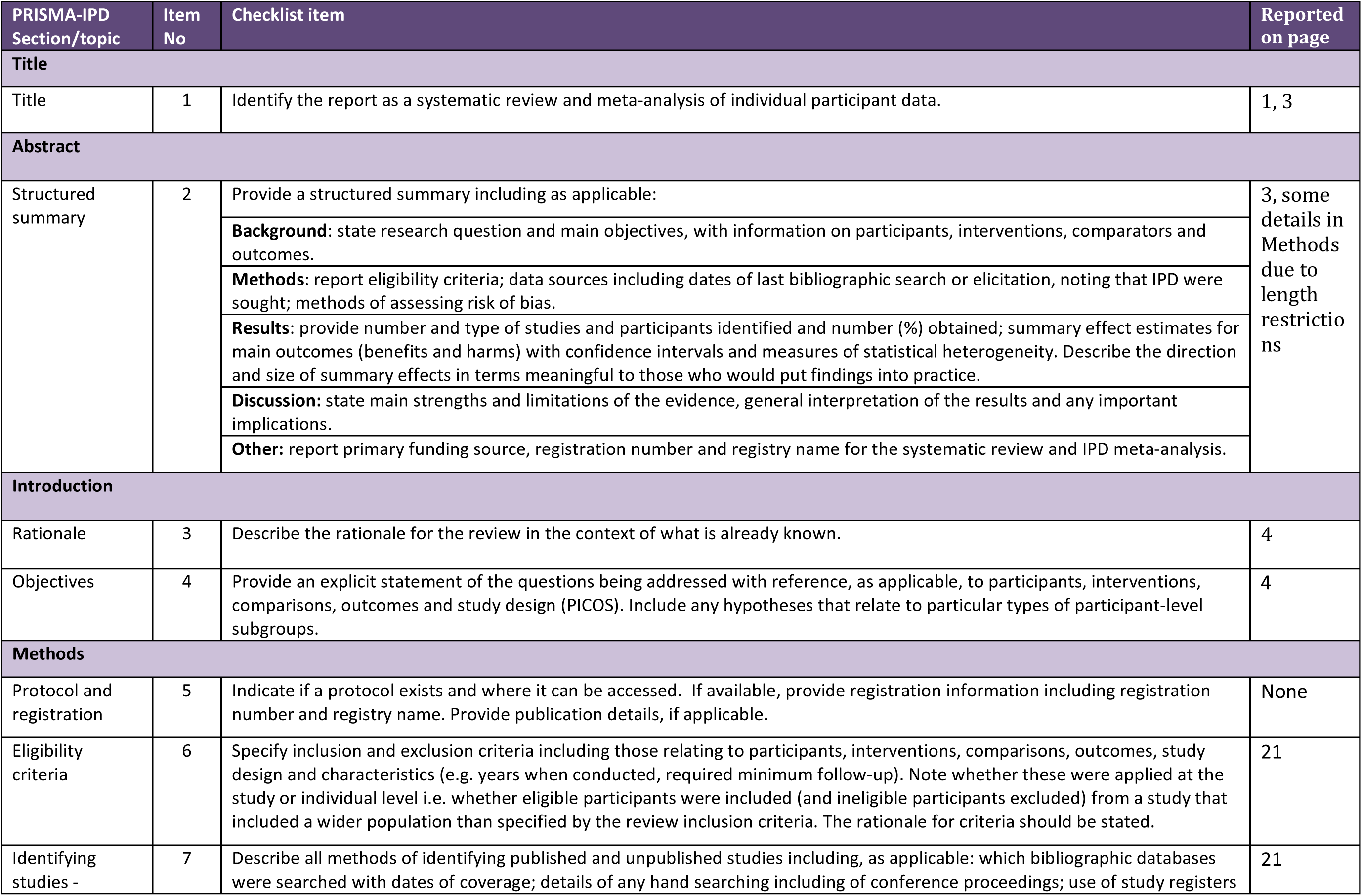
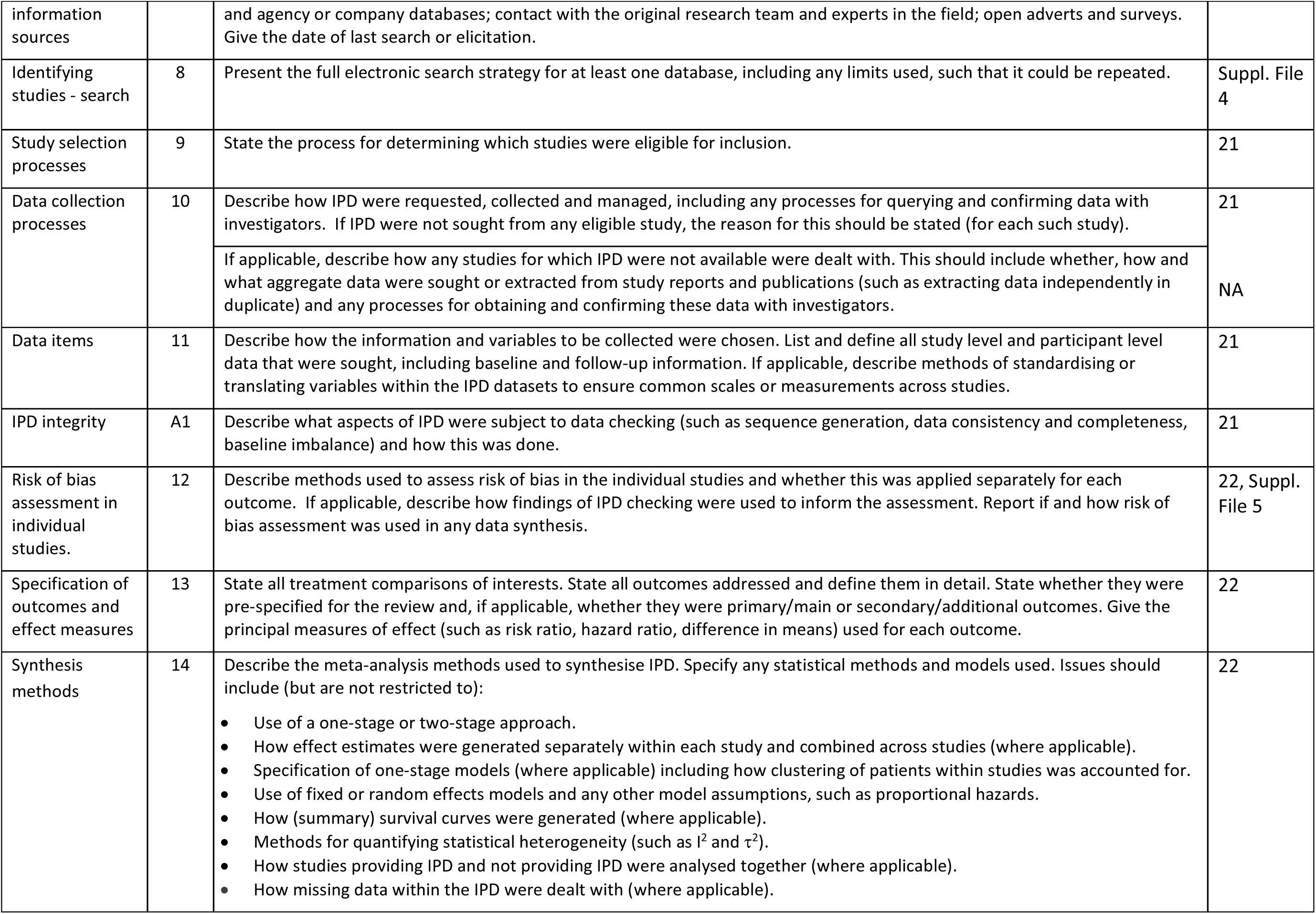
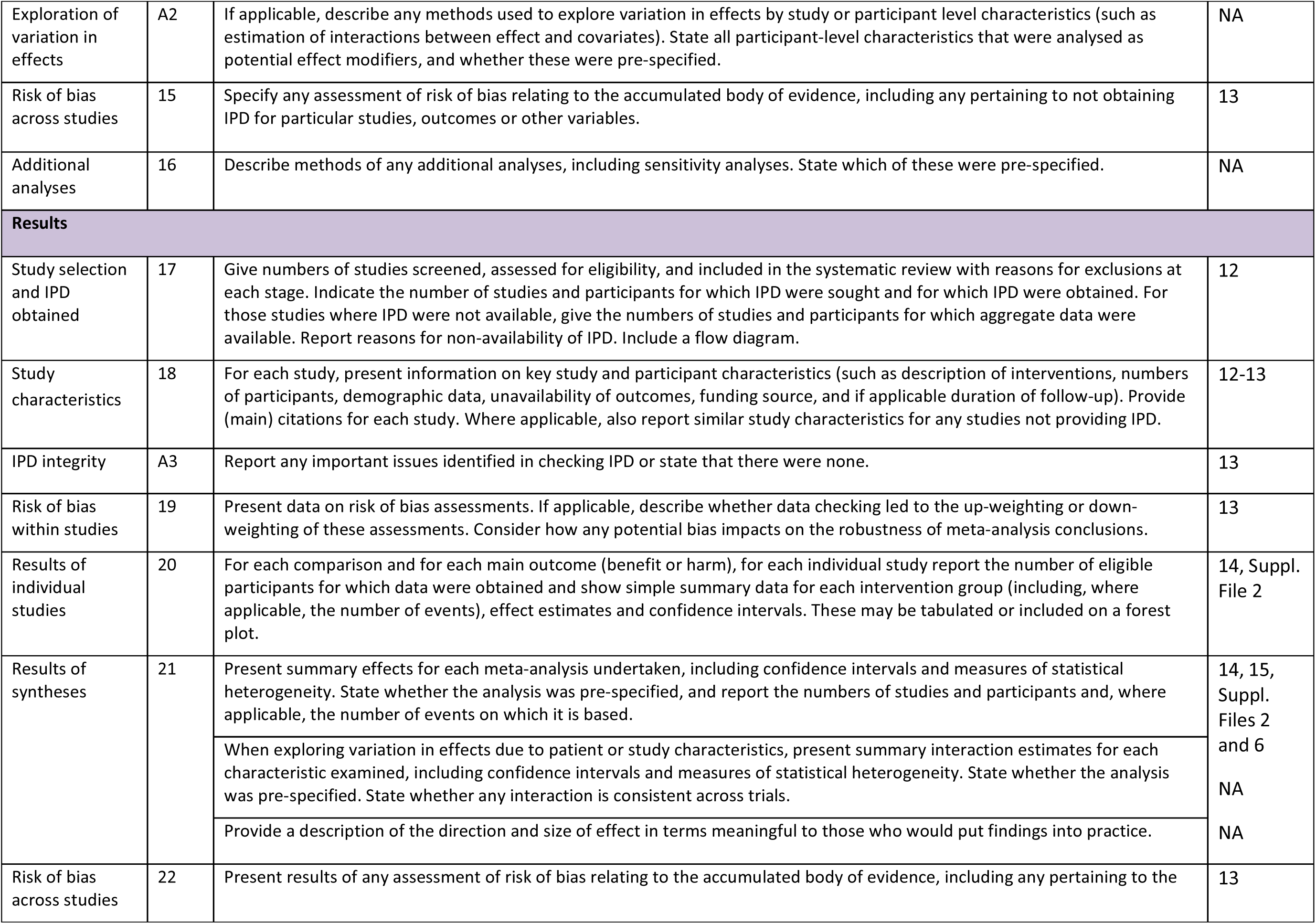
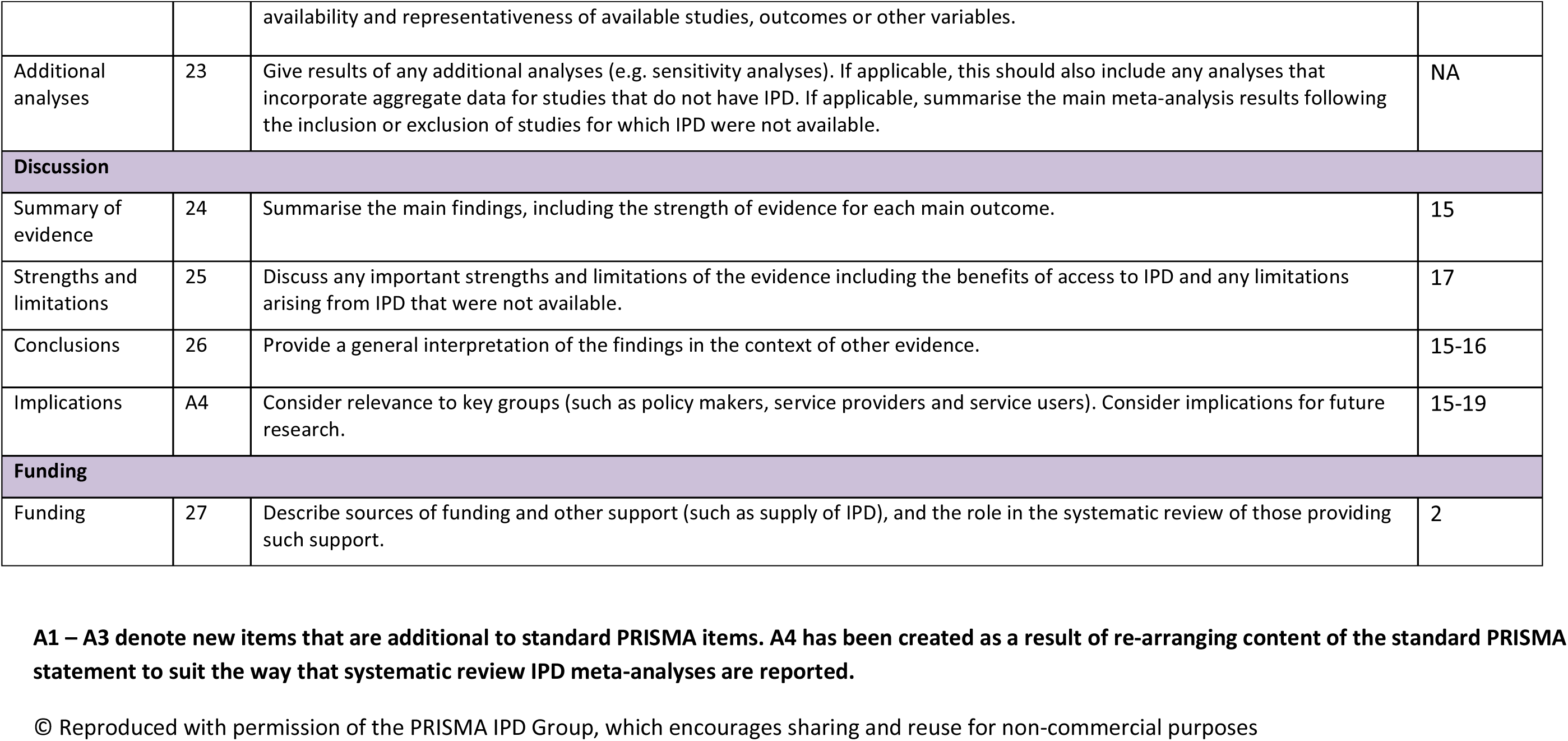

### Supplemental File 4 – Systematic review search terms

(("Immunoproteins"[Mesh] OR "Cytokines"[Mesh] OR "Antimicrobial Cationic Peptides"[Mesh] OR "Immunoassay"[Mesh] OR immunoassay*[tiab] OR cytokine*[tiab] OR interleukin*[tiab] OR immunoprotein*[tiab] OR “immune mediator”[tiab] OR “immune mediators”[tiab] OR “immune biomarker”[tiab] OR “immune biomarkers”[tiab] OR “immune modulator”[tiab] OR “immune modulators”[tiab] OR “immune determinants”[tiab] OR “immune environment”[tiab] OR “immune microenvironment”[tiab] OR complement[tiab] OR immunoglobulin*[tiab] OR antibod*[tiab] OR chemokine* OR interferon* OR lymphokine* OR monokine* OR “tumor necrosis factor” OR “tumor necrosis factors” OR “transforming growth factor” OR “transforming growth factors” OR “antimicrobial peptides” OR “antimicrobial peptide” OR “antimicrobial polypeptide” OR “antimicrobial polypeptides” OR defensin OR defensins)

AND

("Vagina"[Mesh] OR "Cervix Uteri"[Mesh] OR vagina*[tiab] OR cervicovaginal[tiab] OR “cervico vaginal”[tiab] OR cervix[tiab] OR cervical[tiab] OR endocervi*[tiab] OR ectocervi*[tiab] OR softcup[tiab] OR “weck cel”)

AND

(“sexually inactive”[tiab] OR “sexual debut*”[tiab] OR “first sex*”[tiab] OR “first intercourse”[tiab] OR “initiating sex*”[tiab] OR “sexual initiation”[tiab] OR “initiate sex*”[tiab] OR virgin[tiab] OR virgins[tiab] OR virginal[tiab] OR “sexually inexperienced”[tiab] OR “sexually abstinent”[tiab] OR “became sexually active”[tiab] OR “never sexually active”[tiab] OR “no history of sexual intercourse”[tiab] OR “never having had sex”[tiab]))

NOT

(“animals”[mh] NOT “humans”[mh])

### Supplemental File 5 – Risk of bias assessment scale

**MODIFIED NEWCASTLE - OTTAWA QUALITY ASSESSMENT SCALE**

**Risk of bias assessment of pre/post-first sexual intercourse studies**

<u>Note</u>: A study can be awarded a maximum of one star for each numbered item within the Selection and Outcome categories. A maximum of two stars can be given for Comparability

#### Selection

1. Representativeness of the pre-first sex cohort

a. truly representative of the average AGYW in the community ↓
b. somewhat representative of the average AGYW in the community ↓
c. selected group of users eg nurses, volunteers
d. no description of the derivation of the pre-first sex cohort
2. Selection of the post-first sex cohort

a. drawn from the same community as the exposed cohort ↓
b. drawn from a different source
c. no description of the derivation of the post-first sex cohort
3. Ascertainment of pre/post-first sex status

a. self report plus markers of sexual exposure (PSA, y-chromosome, pregnancy, STI, or similar) ↓
b. self report alone
c. no description

#### Comparability

1. Comparability of cohorts on the basis of the design or analysis

a. study controls for bacterial vaginosis ↓
b. study controls for age ↓
c. study controls for any additional relevant factor ↓

#### Outcome

1. Method of measurement of immune mediator concentrations

a. same method used for both cohorts ↓
b. different methods used for each cohort
c. no description
2. Outcome measurements are available for all participants

a. yes ↓
b. no but exclusions are unlikely to introduce bias ↓
c. no and exclusions may introduce bias
d. no description

### Supplemental File 6: Meta-analyses for each immune factor

#### Log2-fold difference between pre and post-first sex

This section shows the meta-analyses for each immune factor where the concentrations of at least half of the samples fell above the limit of detection. A different immune factor is shown on each page.

Each row represents a different study, with the squares indicating the mean and the lines indicating the 95% confidence intervals. Positive numbers indicate higher concentrations post-first sex, while negative numbers indicate higher pre-first sex. The size of the squares is proportional to how heavily the study is weighted in the meta-analysis.

The center of the diamond and the vertical dotted line indicates the meta-effect as determined by the random effects model. The width of the diamond indicates the 95% confidence interval of the meta-effect.

TE, treatment effect (log2-pg/mL of the post-first sex minus log2-pg/mL of the pre-first sex samples); seTE, standard error of the treatment effect; 95%-CI, 95% confidence interval around the treatment effect; Weight, the percentage of the meta-estimate contributed by each study.

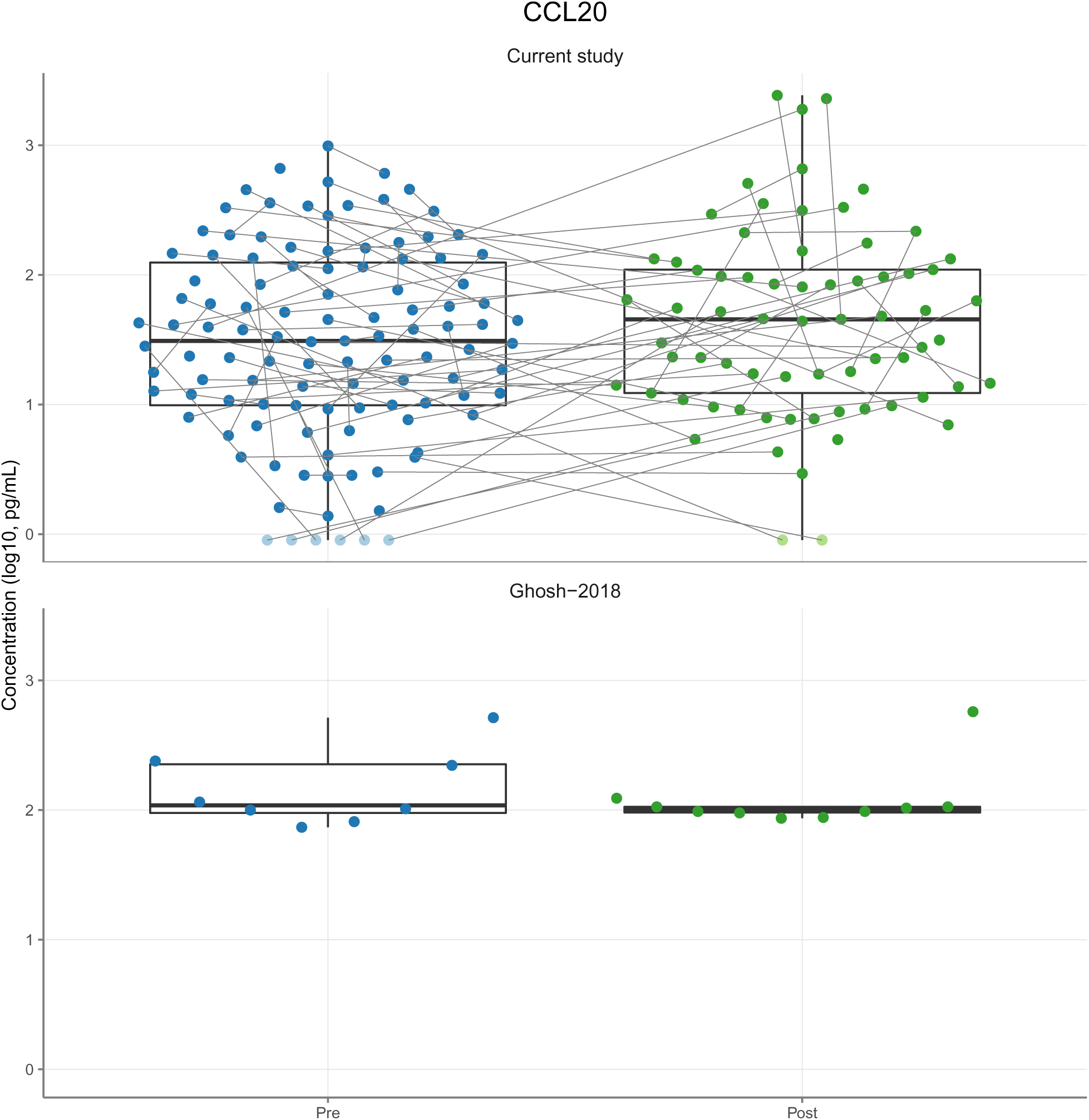
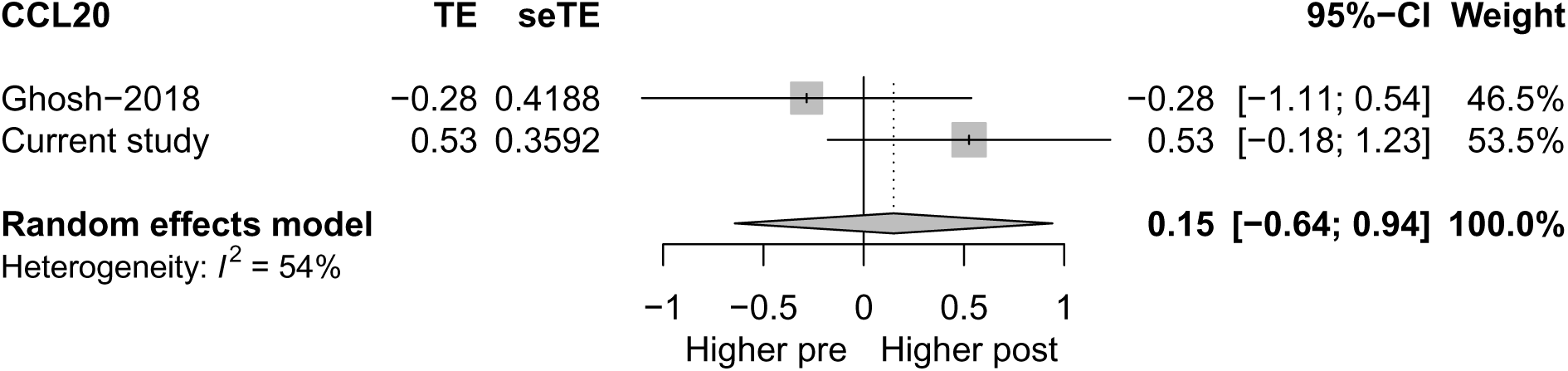
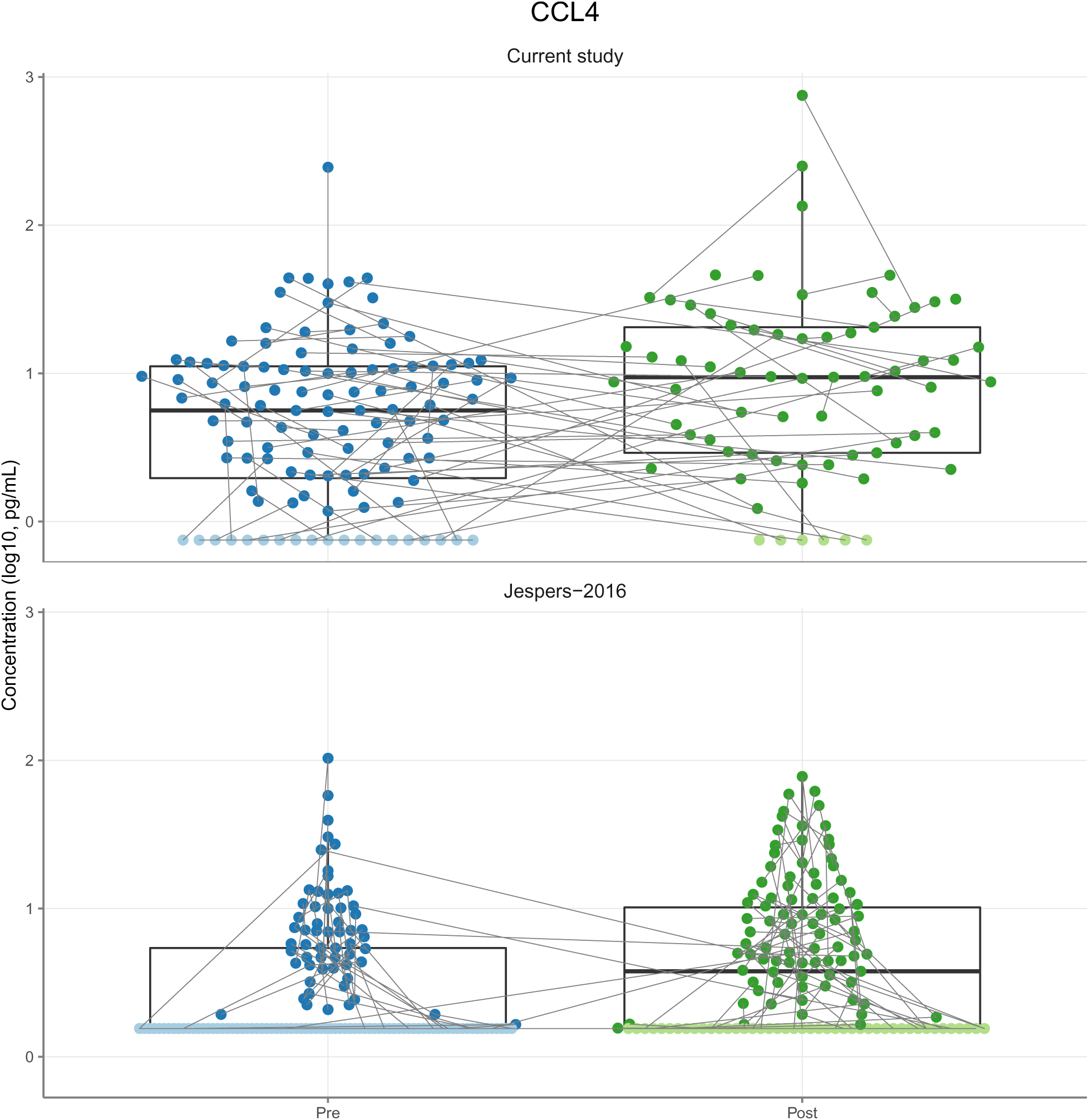
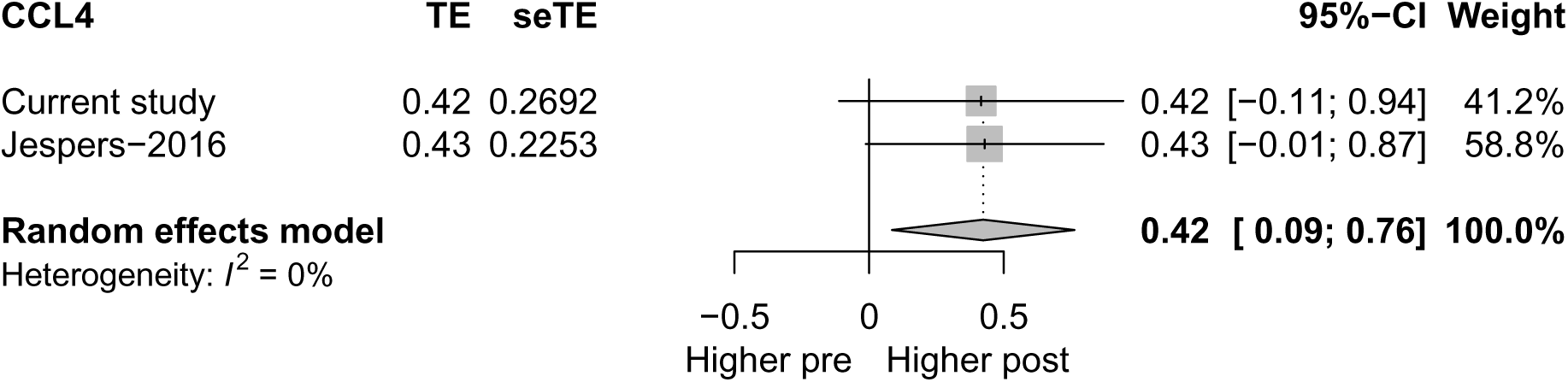
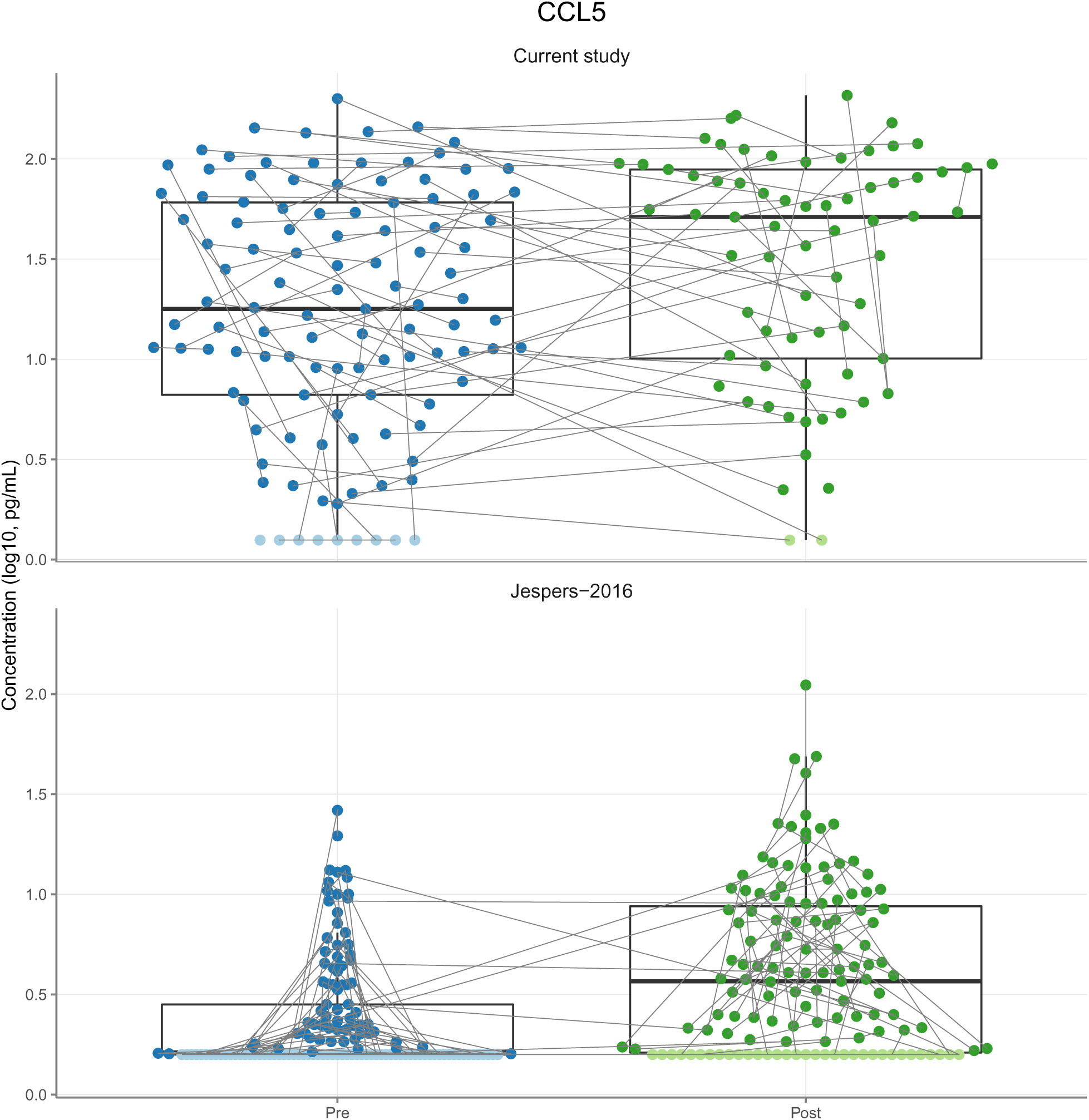
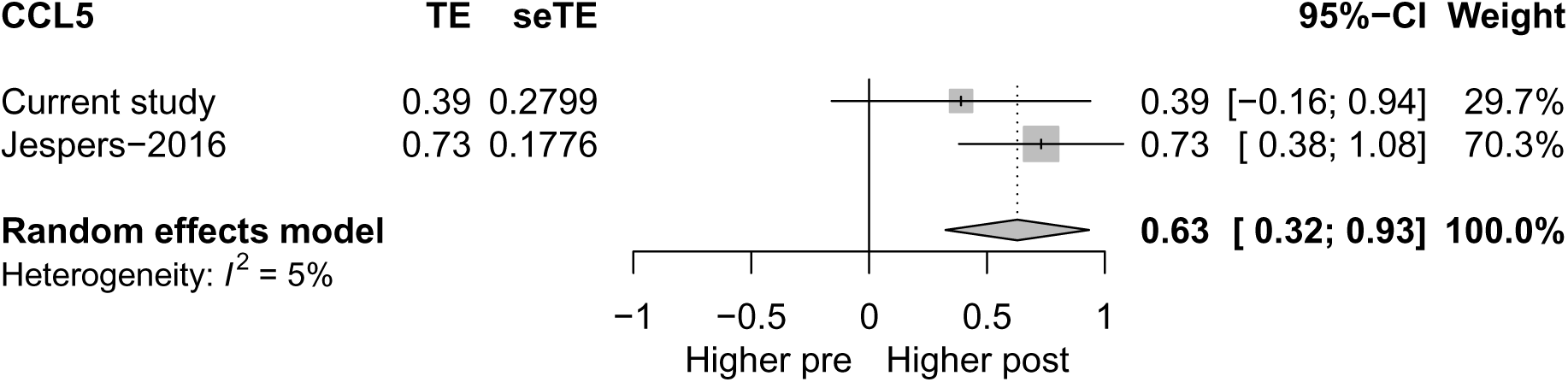
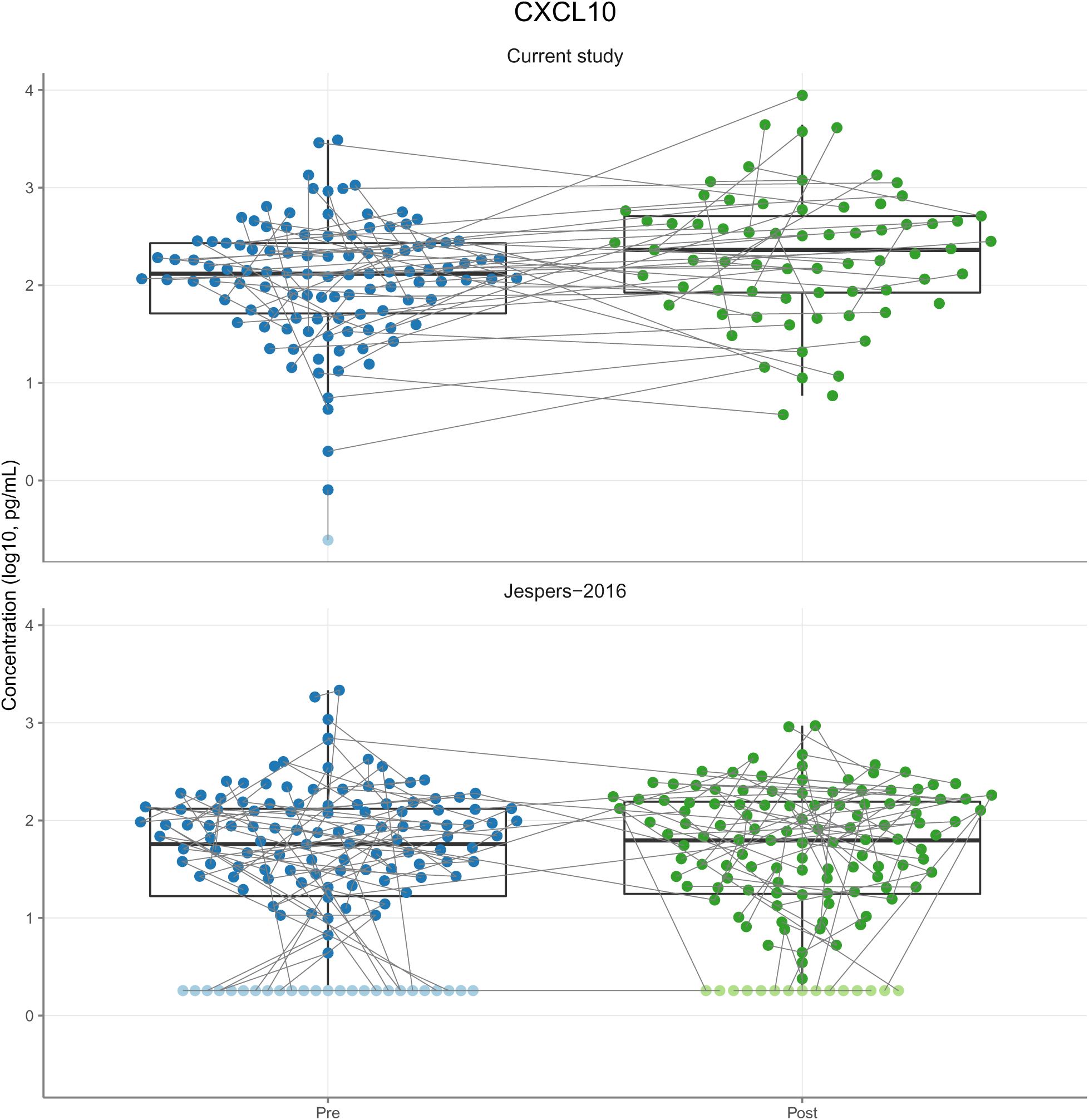
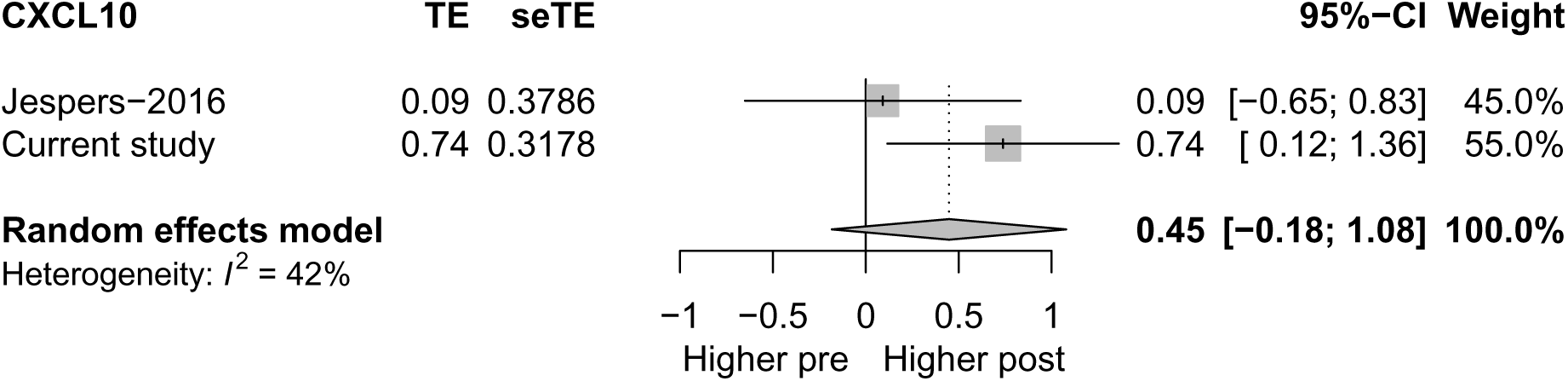
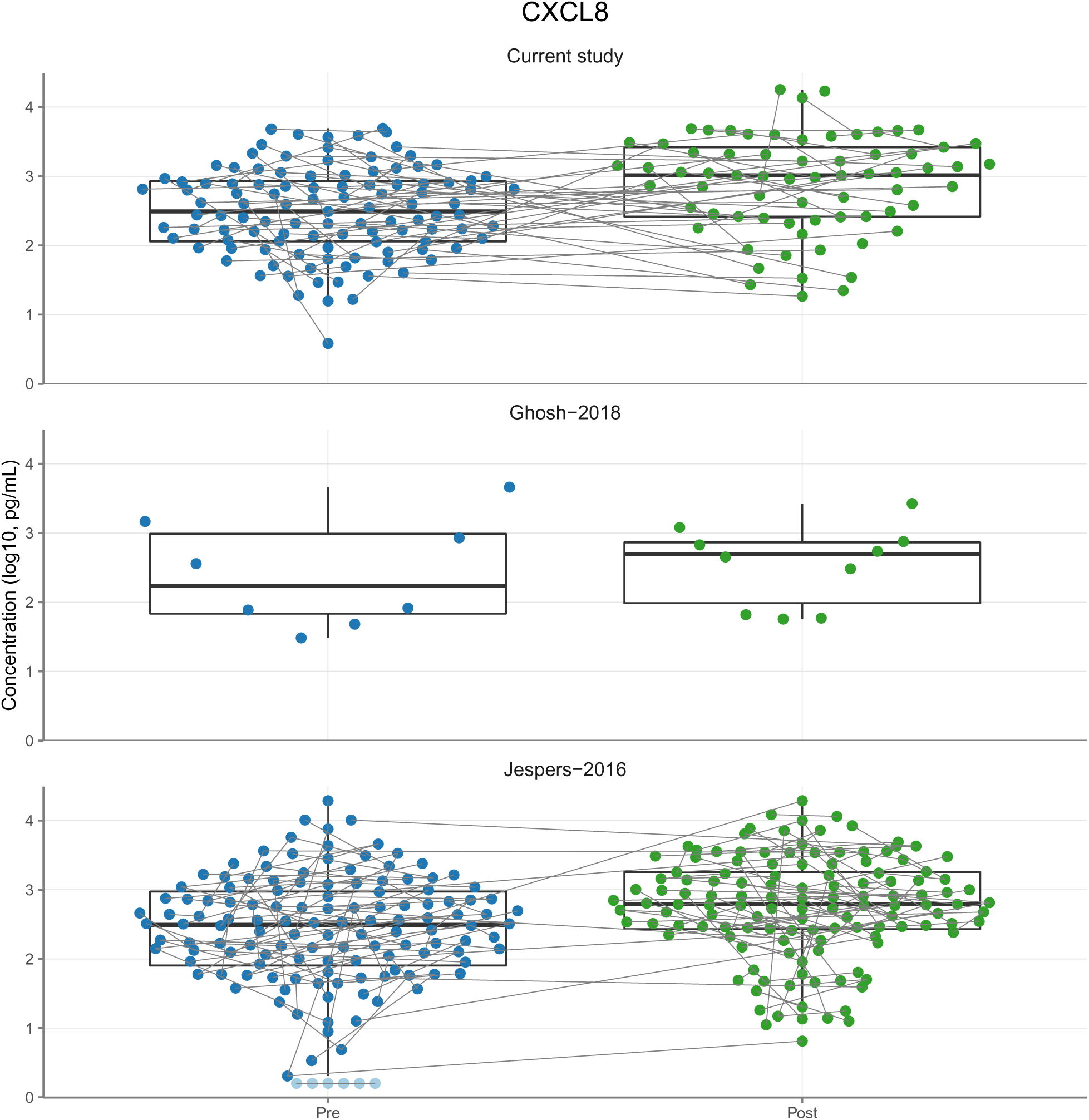
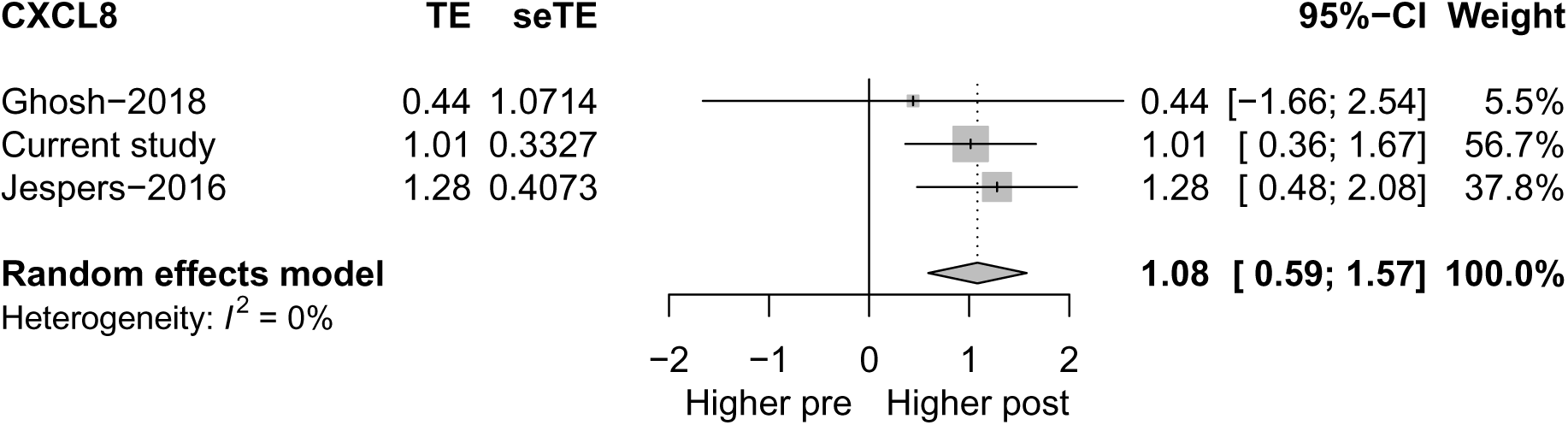
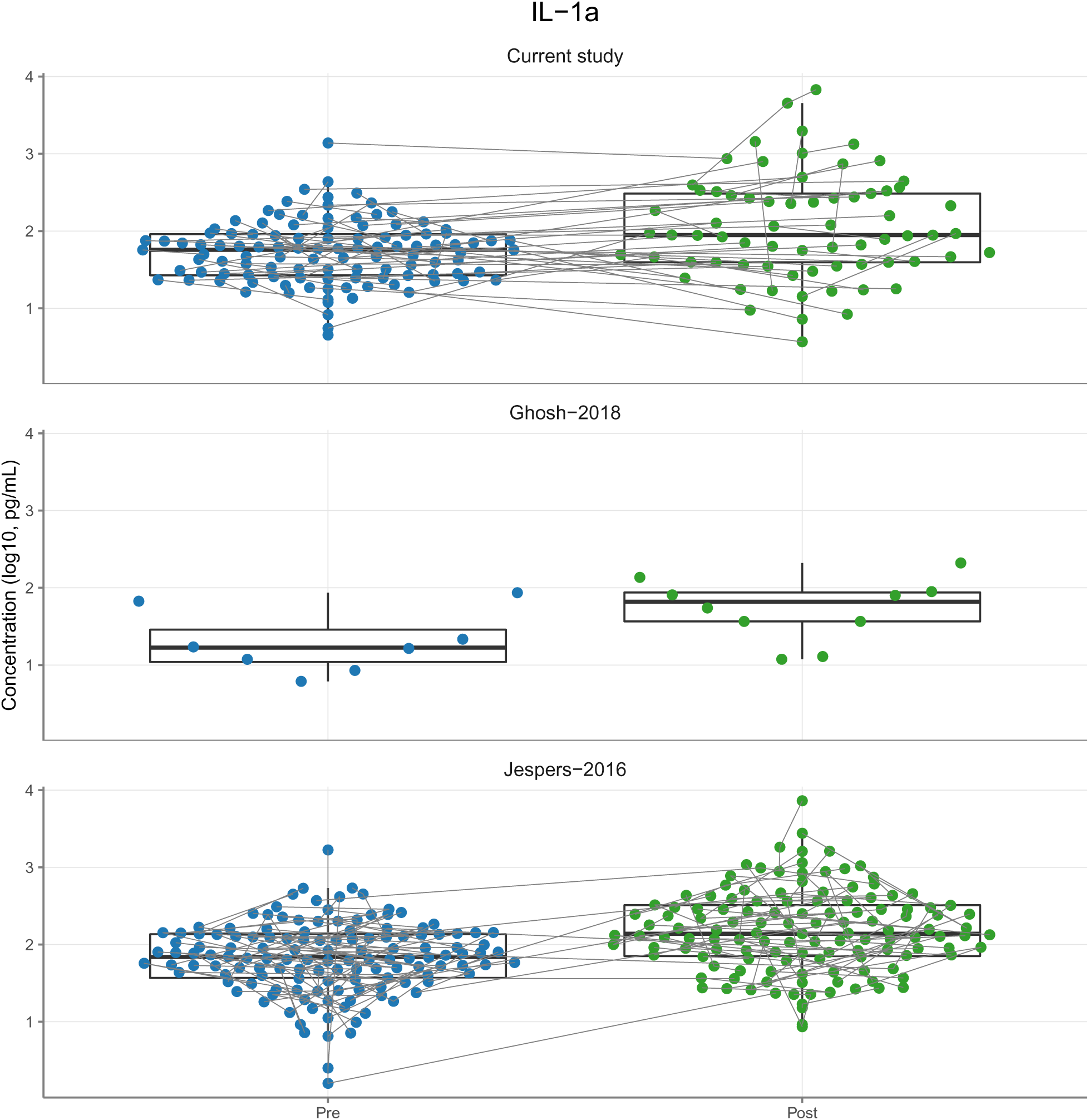
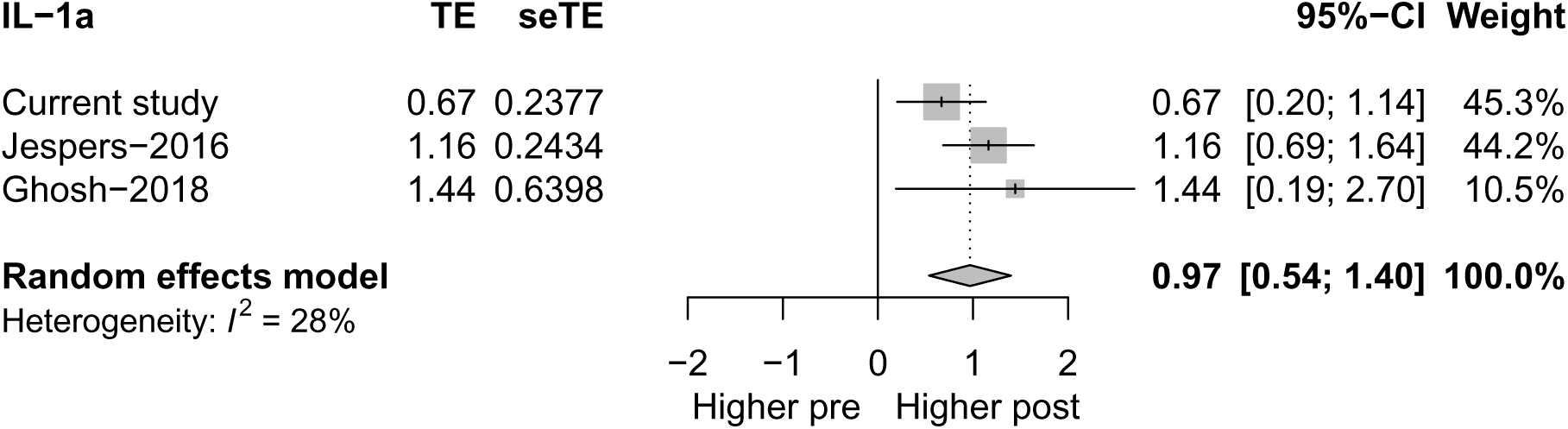
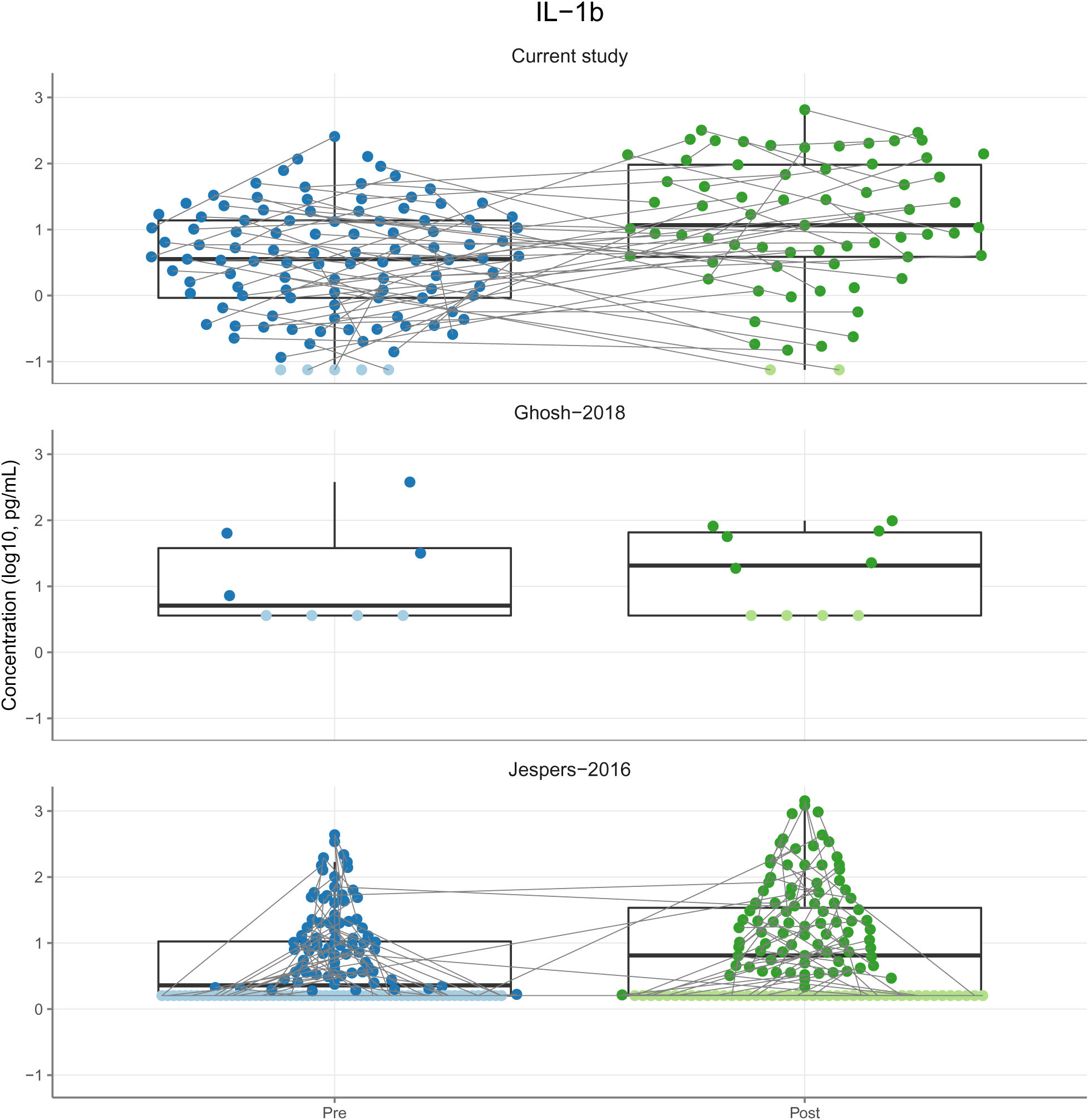
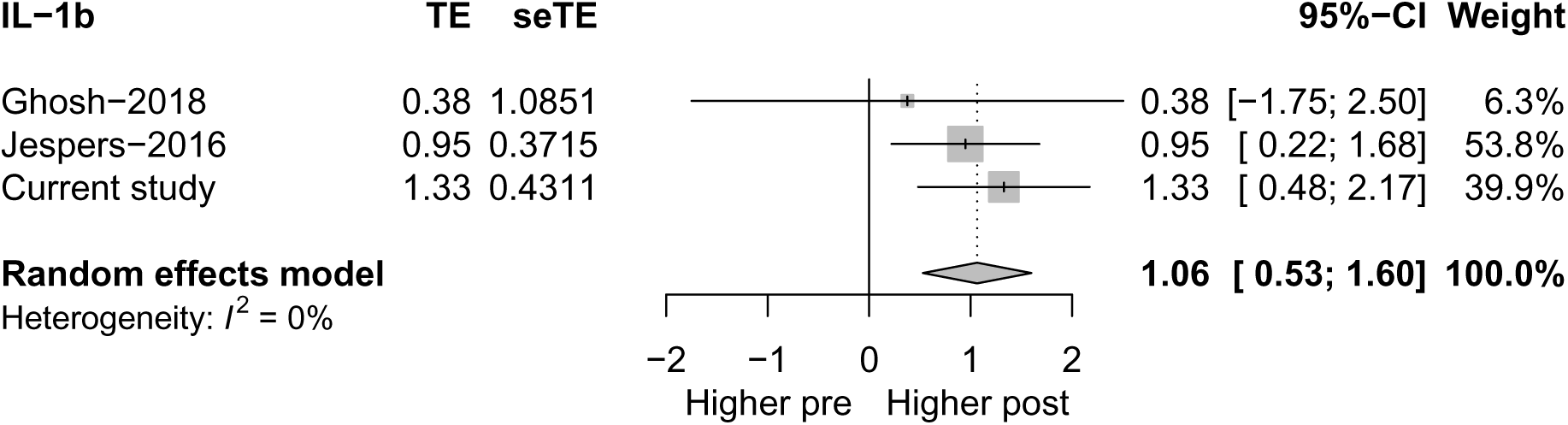
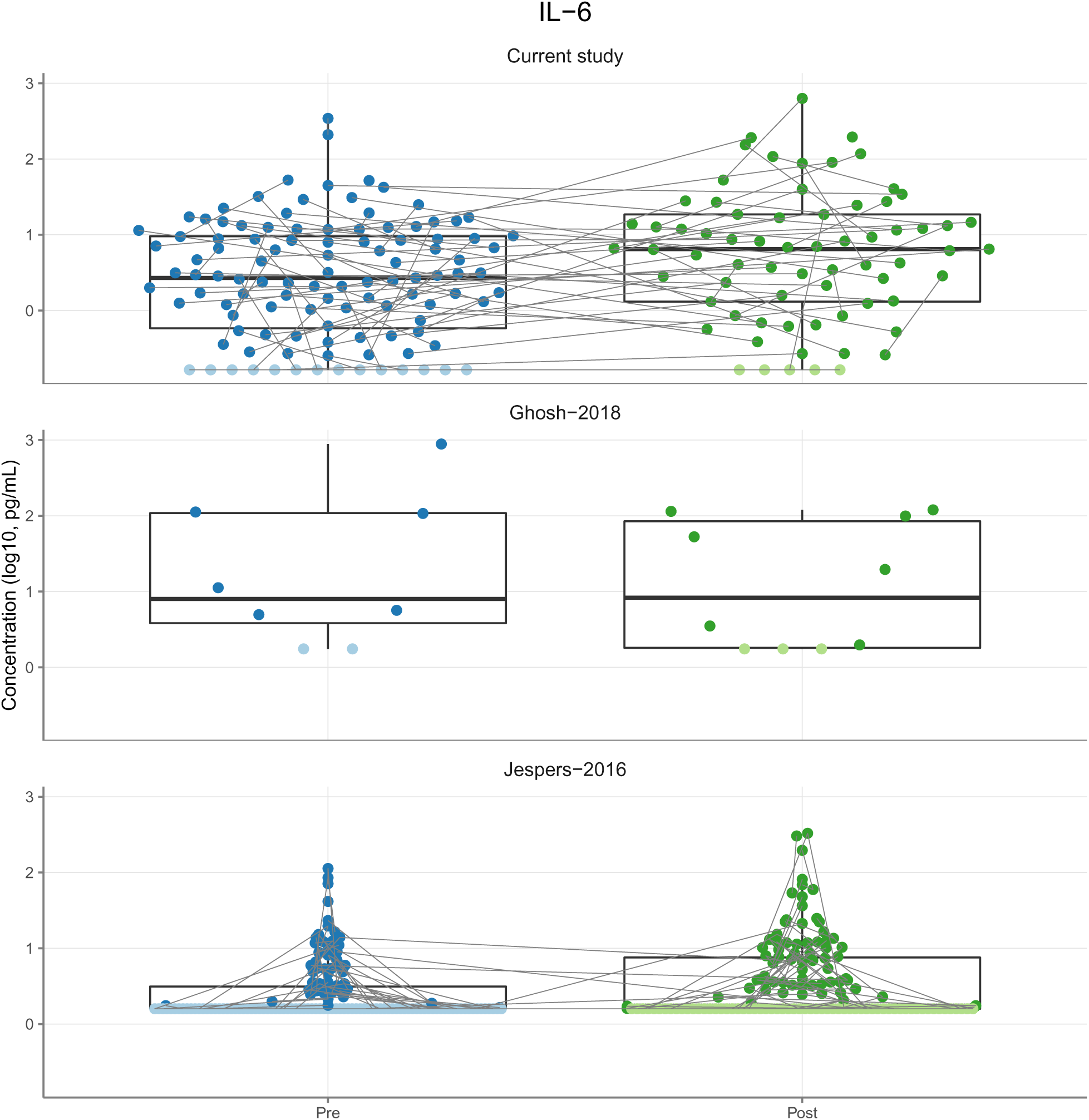
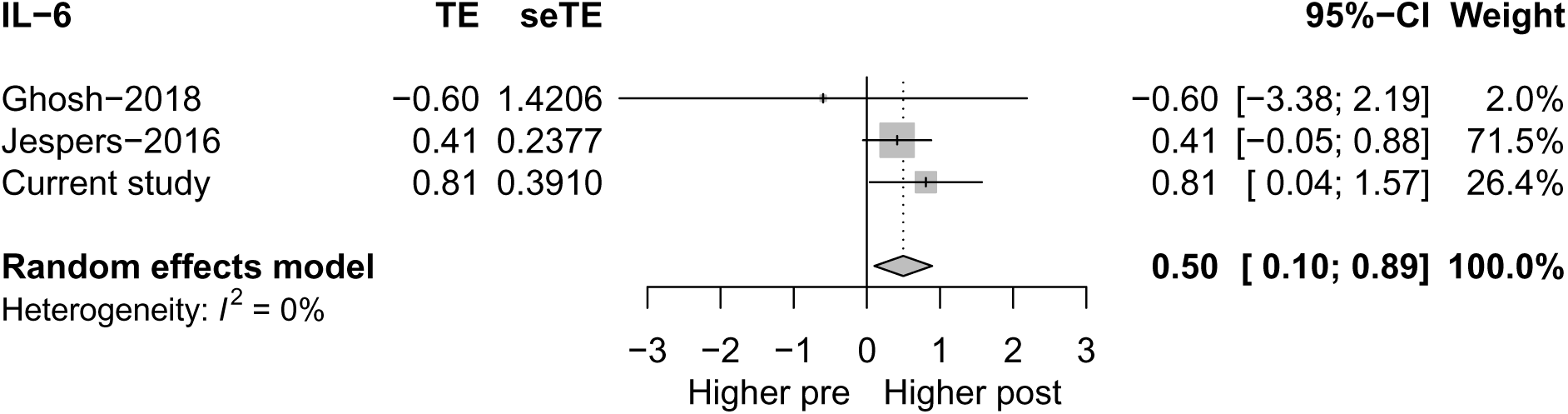
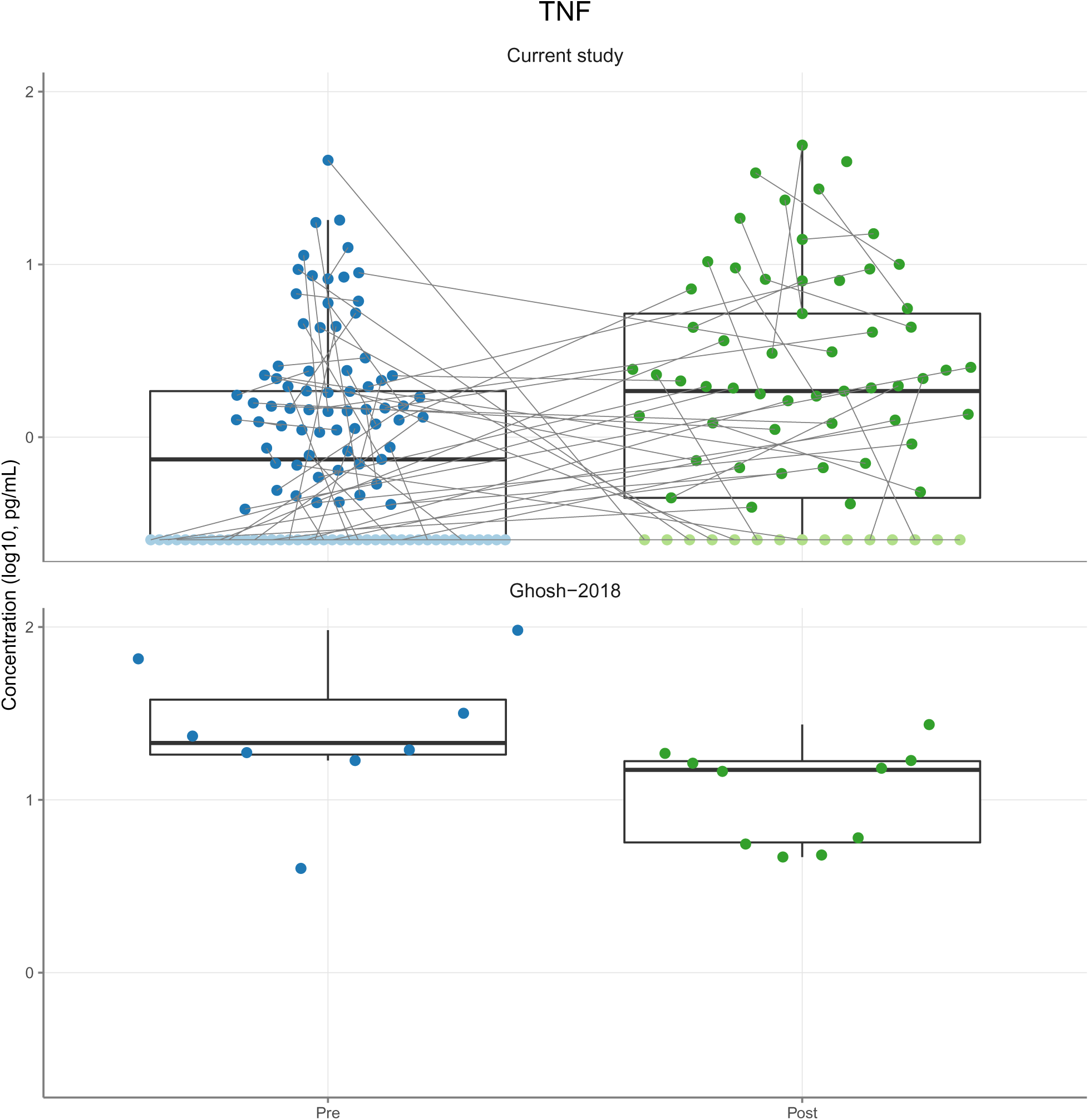
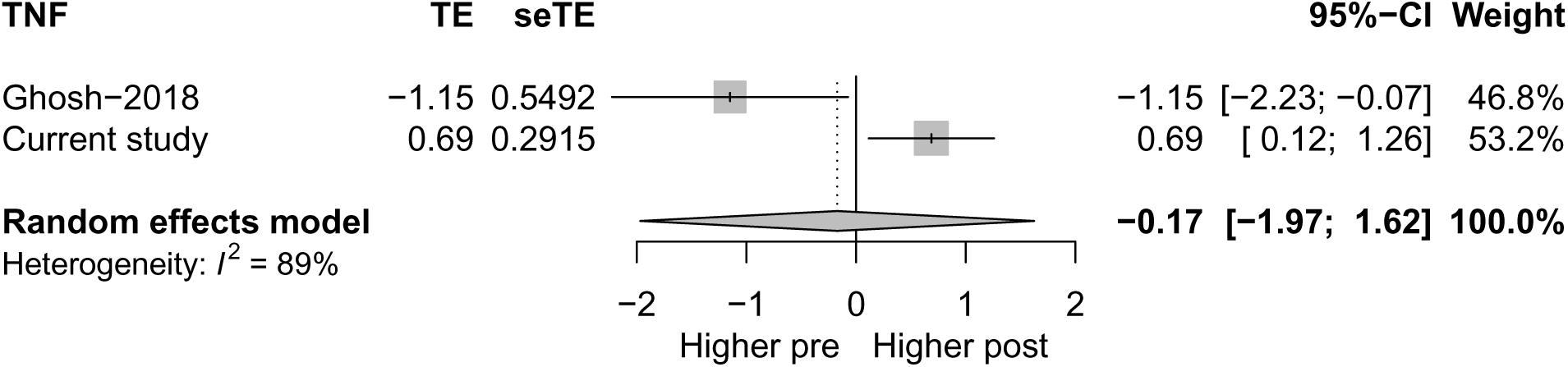

